# Electrochemical immunosensor-enabled liquid biopsy of extracellular vesicles for diagnosis of metabolic dysfunction-associated steatohepatitis

**DOI:** 10.64898/2026.09.02.26362088

**Authors:** Thi Thanh-Qui Nguyen, Daheui Choi, Seonhwa Lee, Jose M. de Hoyos-Vega, P. Vineeth Daniel, Amy S. Mauer, Michael J Eller, Anthony Giron, Diana F. Cedillo-Alcantar, Gulnaz Stybayeva, Rondell P. Graham, Alina M. Allen, Konstantinos N. Lazaridis, Harmeet Malhi, Alexander Revzin

**Author notes:** These authors contributed equally to this work as first author (Thi Thanh-Qui Nguyen, Daheui Choi).

## Abstract

Accurate, noninvasive biomarkers for diagnosis and longitudinal monitoring of metabolic dysfunction–associated steatohepatitis (MASH) remain an urgent unmet clinical need. Here, we describe an electrochemical immunoassay that involves capturing hepatic extracellular vesicles (EVs) on antibody-modified electrodes followed by detecting EV-borne biomarkers using redox active gold nanoparticles (AuNPs). The intensity of electrochemical redox signal represents a quantitative measure of biomarker abundance. Three MASH-associated biomarkers—S100A11, TNF-α, and TGF-β1—were analyzed on hepatic EVs to assess steatotic, inflammatory, and fibrotic states of the diseased liver. The electrochemical signals for individual biomarkers were added into a composite EV_MASH_ score. Plasma samples from 28 patients with MASH, 22 healthy controls, and 27 patients with non-MASH liver disease were scored in this manner, revealing 100% accuracy in the training cohort and >90% accuracy in an independent validation cohort. Our electrochemical immunosensor represents a minimally invasive liquid liver biopsy, offering a promising alternative to solid liver biopsies for screening and monitoring of MASH.

**One Sentence Summary:** Circulating hepatic extracellular vesicles carry biomarkers of liver injury enabling accurate, minimally invasive MASH diagnosis in blood.

## INTRODUCTION

The prevalence of obesity has become a significant public health concern, contributing to a range of serious health consequences, including cardiovascular diseases, type 2 diabetes, and metabolic dysfunction (*1, 2*). Metabolic dysfunction-associated steatotic liver disease (MASLD) is a chronic liver condition that has become increasingly prevalent worldwide, affecting over 30% of the population (*3, 4*). MASLD presents a wide spectrum of phenotypes, ranging from non-progressive steatosis to the co-existence of steatosis and inflammation, a condition referred to as metabolic dysfunction-associated steatohepatitis (MASH), which is present in 20-30% of those with MASLD (*5, 6*). MASH represents a more aggressive form of liver disease and is a leading cause of cirrhosis, hepatocellular carcinoma, and the need for liver transplantation (*7–9*). MASH is typically asymptomatic and is diagnosed when incidentally elevated liver tests or steatosis are noted. Though there has been significant progress in non-invasive tests for monitoring patients that have known MASH, liver biopsy remains the clinical gold standard for diagnosis (*10*). However, liver biopsy is invasive, with risk of bleeding, infection and inflammation, is costly, and requires specialized expertise (*11*). Furthermore, imaging-based methods provide only partial information regarding the disease stage or its severity and are hampered by cost and scalability (*10*). Noninvasive scores, such as FIB-4, are impacted by age and disease prevalence. Therefore, alternative approaches are needed to address these limitations, offering real-time monitoring of the disease stage, allowing convenient repeated testing, and even estimates of disease progression with minimum difficulty.

Extracellular vesicles (EVs) are nanometer-sized, membrane-bound particles (50-1,000 nm in diameter) secreted by cells (*12, 13*). They are present in bodily fluids, such as saliva, serum, or urine and function as mediators of intercellular communication (*14, 15*). Due to their ability to carry biological information of cells of origin, EVs have been investigated as diagnostic biomarkers (*15–18*). In the context of MASH-associated EVs, several studies have identified that circulating EVs are present in significantly greater numbers in plasma samples from patients with chronic liver diseases compared to healthy controls (*19–21*). Other studies reported that the numbers of hepatic EVs and expression of hepatic markers (e.g. Cytochrome P450 2E1; CYP2E1 and Asialoglycoprotein Receptor 2; ASGR2) on these EVs decreased after bariatric surgery in patients with MASLD (*2, 18*). There is therefore evidence for EV numbers and composition being reflective of liver disease status. However, to the best of knowledge, there has not been a report connecting the composition of circulating EVs to the steatotic, inflammatory or fibrotic state of the liver. Our study represents a first step in the direction of a liquid biopsy being used to stage and grade the severity of disease in MASH patients.

A number of analytical approaches has been developed for EV characterization, including immunofluorescence (*2, 22, 23*), colorimetric (*24–27*), mass spectroscopy (*28*), surface plasmon resonance (SPR) (*29–32*), and electrochemical analysis (*15, 23, 33, 34*). All of these approaches have merits and contribute to the overall understanding of EV composition. Our ultimate goal is to develop a cost-effective and user-friendly assay suitable for the clinical setting. Electrochemical platforms are particularly attractive in this context due to their high analytical sensitivity, scalability, and low instrumentation cost. There have been several examples of electrochemical immunoassays applied toward EV analysis. In one demonstration, nano-interdigitated electrodes were used to detect cancer EVs (*35*). In another example, a microfluidic chip with electrochemical biosensors was used to quantify EVs in patients with pancreatic cystic neoplasms (*36*). In yet another example, an integrated magneto-electrochemical device was developed for isolating EVs and profiling colorectal cancer-related protein markers from plasma (*34*).

Our team has developed an electrochemical microtiter plate integrating microfluidic capillary valves and an array of electrodes to perform sensitive electrochemical immunoassays in a multi-well format (*15*). In this study, we sought to leverage this system for analysis of MASH-associated biomarkers on hepatic EVs. We first validated the platform using EVs derived from injured hepatocytes and then proceeded with testing 77 patient plasma samples. The electrochemical immunosensor had excellent sensitivity and specificity in distinguishing MASH samples from other liver diseases and healthy controls. The diagnostic platform described here represents a liquid liver biopsy that is point-of-care deployable and may enable future screening of patients for MASH and other liver diseases.

## RESULTS

### Characterization of Lipotoxicity in Primary Human Hepatocytes

In this study, we chose to focus on — Transforming Growth Factor-beta 1 (TGF-β1), Tumor Necrosis Factor-alpha (TNF-α), and S100 calcium-binding protein A11 (S100A11) as markers of fibrosis, inflammation and lipotoxicity, respectively. Our team and others in the field have connected S100A11, a calcium binding protein, to endoplasmic reticulum (ER) stress associated with lipotoxic injury in MASH (*37, 38*). This led us to hypothesize that S100A11 may be present on hepatic EVs and may represent a biomarker of lipotoxicity. Similarly, TGF-β1 and TNF-α are well established as markers of liver fibrosis and inflammation, respectively (*39–41*). Therefore, we reasoned that these proteins may be present on EVs originating from the injured liver. Given that liver-derived EVs represent a small fraction (<1%) of the total circulating EV population (*42*), we wanted to implement an enrichment step as part of our assay. Building on previous findings that hepatic EVs express hepatocyte-specific surface markers, including ASGR2, (*2, 28*) we leveraged this marker to selectively enrich hepatic EVs for downstream analysis.

With these considerations in mind, we proceeded to develop an assay for enriching hepatic EVs on a sensing surface and then detecting the presence of injury markers on the EVs. To conserve precious patient plasma samples, we used an *in vitro* liver injury model to establish and validate our assay. The primary human hepatocytes isolated from a chimeric mouse model (PXB-mice®) (*43*) were cultured (Figure 1A) exposed to palmitic acid (PA) to simulate lipotoxic hepatocyte injury, a cardinal feature of steatohepatitis (Figure 1B) (*44*). Based on the reported pathophysiological levels of PA, we induced lipotoxic injury in hepatocytes by exposing cells to 0.4 mM PA, thereby promoting lipid accumulation and EV secretion. PA treatment led to 10.87-fold greater intracellular lipid droplet accumulation than untreated cells (Figure 1C). Immunofluorescence staining revealed higher levels of TGF-β1 (fibrosis marker), TNF-α (inflammation marker) and S100A11 (steatosis marker) in PA-treated hepatocytes (see Figure 1D), while albumin (ALB) expression remained unchanged (Figure 1E). These findings were consistent with PCR analysis which showed stable expression of hepatocyte identity genes (ALB and CYP2E1) alongside upregulation of the three inflammation- and injury-related genes in PA-treated cells (Figure 1F). Collectively, these results demonstrate that lipotoxic injury induces upregulated expression of biomarkers associated with steatosis, inflammation, and fibrosis at the cellular level (*45, 46*).

**Figure 1.**
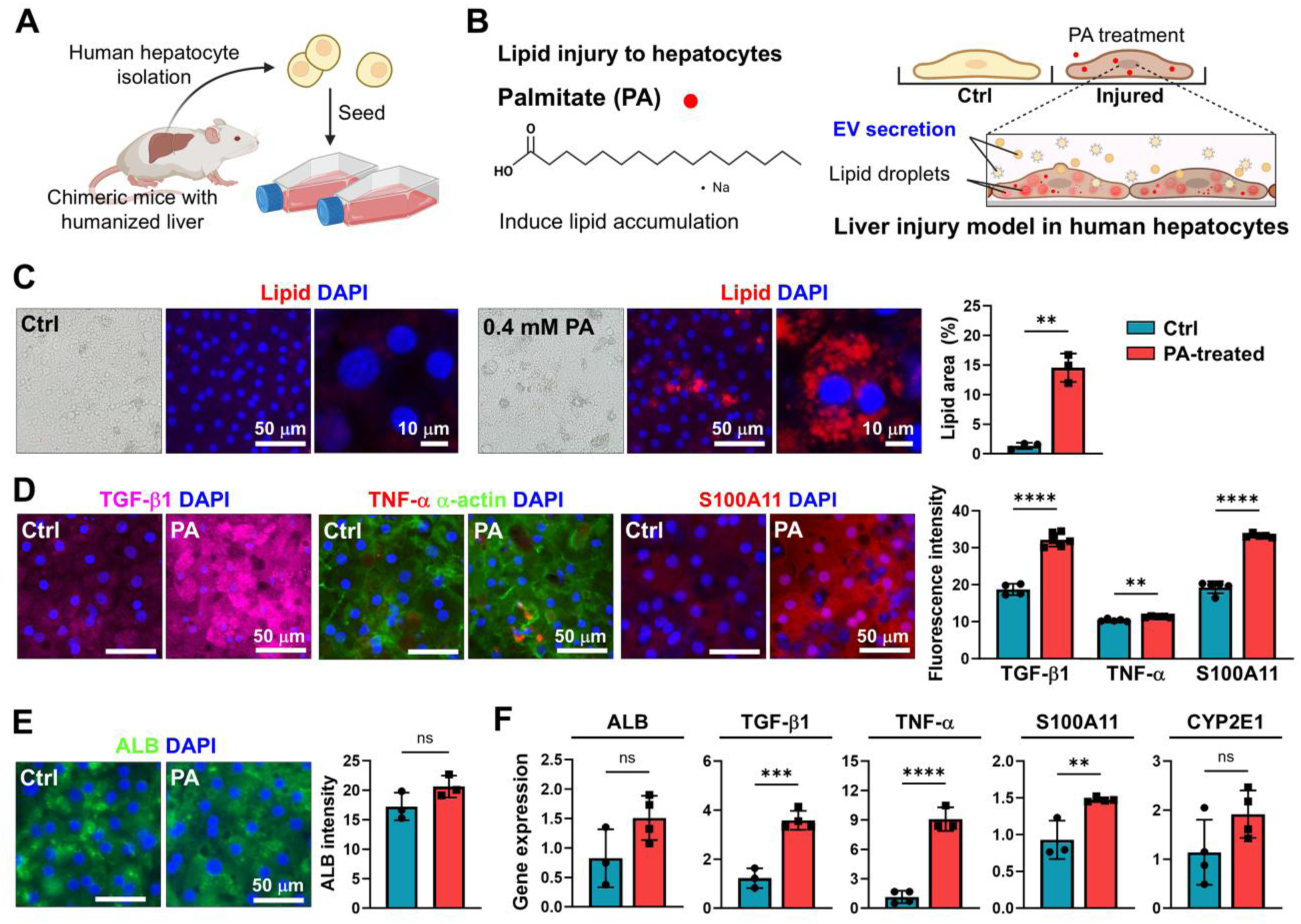
Assessing lipotoxic injury model using primary human hepatocytes. (A and B) Illustration of the experimental workflow. The human hepatocytes isolated from mice with humanized liver were seeded on 75 T-flasks and then lipotoxic injury was initiated by exposure to palmitate (PA). (C) Brightfield and lipid droplet accumulation image of human hepatocytes at 72 h, with and without PA exposure. Immunofluorescence staining of (D) TGF-β1, TNF-α and S100A11 and (E) albumin (ALB) in controls and PA-injured hepatic cell after 72 h. The quantification of fluorescence signals associated with lipid accumulation and immunofluorescence staining is presented in the bar graph. (F) RT-PCR analysis of biomarker gene expression in control and PA-treated hepatic cell spheroid (ns p>0.05; * p<0.05; ** p>0.01; *** p<0.001; **** p<0.0001).

### Characterization of EV from lipotoxic injured hepatocytes

Given our hypothesis that injury at the cellular level may be reflected in the composition of EVs, we proceeded to harvest EVs from injured and healthy human hepatocyte cultures. For these experiments, cells were maintained in media with EV-depleted serum. EV isolation was carried out by a combination of sucrose-gradient separation and ultracentrifugation (see Figure 2A). EVs were characterized using several parallel methods in accordance with Minimum Information for Studies of Extracellular Vesicles (MISEV) recommendations (*47*). First, isolated EVs were characterized by nanoparticle tracking analysis (NTA) and were confirmed to have an average diameter of 91.2 nm for control and 88.7 nm for PA-treated EVs, which is consistent with small EVs (Figure 2B). Particle concentration was also established by NTA and was observed to be 4.88 times higher for injured hepatic cells compared to controls. This observation was consistent with previous reports by our team and others pointing to higher levels of EV production associated with liver injury (*2, 42*). Next, we functionalized gold (Au) substrates with anti-CD63 antibodies (Abs) and captured EVs on these substrates. Scanning electron microscopy (SEM) analysis revealed the presence of spherical particles consistent in size and shape with EVs (see Figure 2C). In contrast, substrates functionalized with isotype control (rabbit IgG) Abs showed negligible particle binding, confirming the specificity of EV capture on Au substrates (Figure S1). As the next step in the MISEV-recommended EV benchmarking protocol, western blot analysis was used to evaluate the presence of ubiquitous EV markers including TSG101, CD9 and Syntenin. As seen from Figure 2D, particles isolated from media conditioned by both normal and injured hepatic cells expressed characteristic markers of EVs.

**Figure 2.**
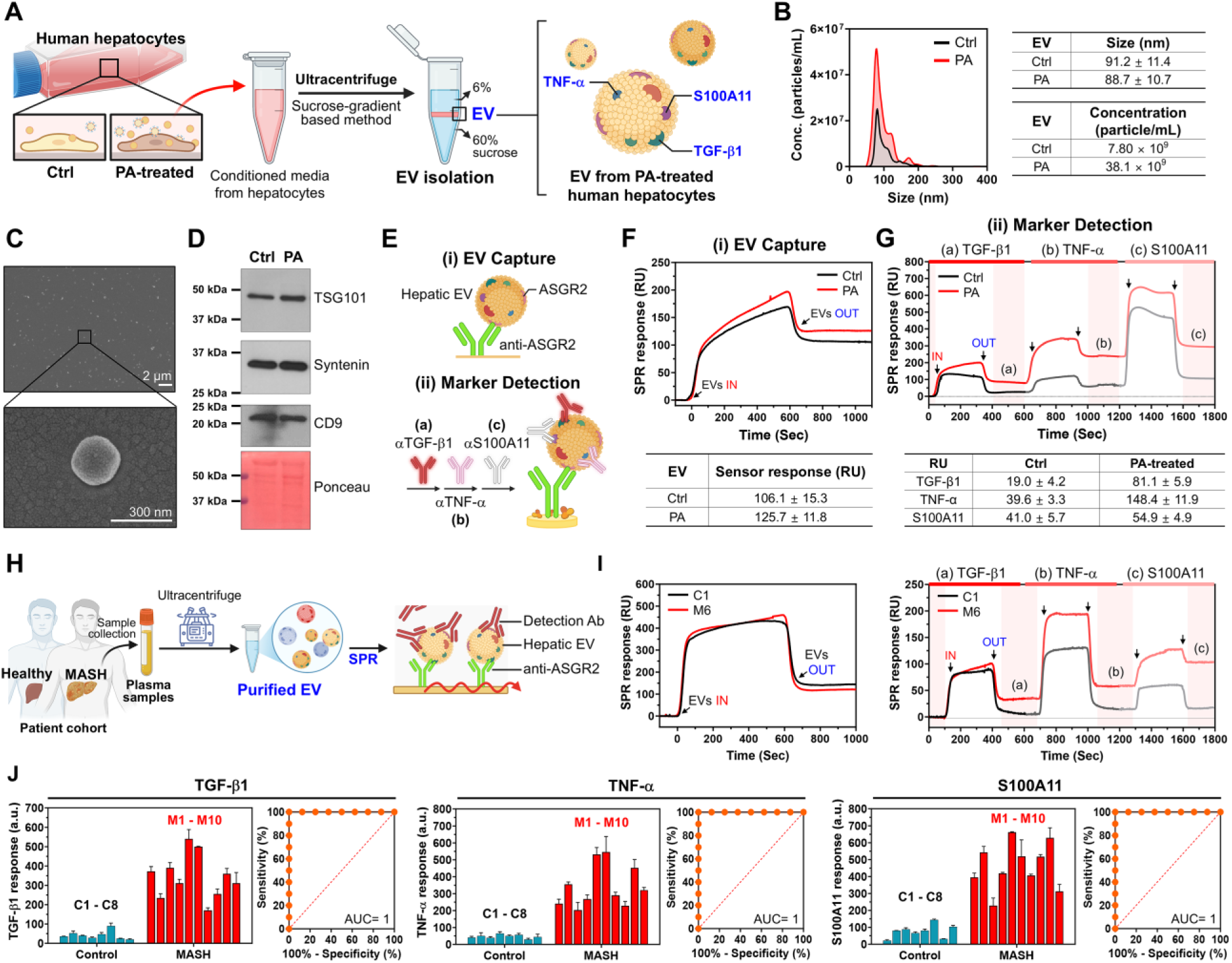
Characterization of injured hepatic EV from hepatocytes and patient plasma. (A) Scheme of EV isolation from cell-conditioned media by density gradient separation method using sucrose. Isolated EVs were analyzed for expression of TGF-β1, TNF-α and S100A11. (B) NTA size distribution and concentrations of EVs isolated from control (Ctrl) and PA-treated human hepatocytes. (C) SEM images of EVs captured on an Au substrate functionalized with anti-CD63. (D) Western blot analysis of EVs from Ctrl or PA-treated spheroids. An equal volume of 20 µL EV suspension (approximately 5 × 10¹¹ particles/mL) was loaded per lane. Representative blots for Tsg101, Syntenin, and CD9 are depicted. Ponceau red staining of the membrane was employed as a loading control. (E) Illustration on principal of SPR steps including (i) EV capture; the step for capturing hepatic specific EVs on anti-ASGR2-immobilized surface and then (ii) Marker detection; the step for sequentially labeling 3 Abs and measuring expression level of markers on EVs. (F) SPR analysis comparing binding of EVs on Au substrates functionalized with anti-ASGR2. Binding of EVs from injured and control hepatocytes was compared. (G) SPR binding profile of EVs from PA treated and untreated cultures were captured on anti-ASGR2 substrate and analyzed for expression of anti-TGF-β1, anti-TNF-α and anti-S100A11. Labeling sequence shown in the cartoon was found to be optimal. (H) Illustration of the experimental workflow. Healthy control and MASH plasma sample cohorts were analyzed. EVs were isolated from each plasma sample, adjusted to parity between plasma samples, captured on SPR chips using anti-ASGR2 and analyzed for expression of putative biomarkers. (I) Comparison for EV binding for chosen control and MASH samples. Note similar levels of binding for MASH and control EVs, indicating that ASGR2 expression levels are not affected by disease/injury. SPR analysis of Ab labeling experiment for control and MASH EVs captured on anti-ASGR2 surface. Note that binding signals for all three biomarkers are much higher on EVs from MASH sample. (J) Summary of TGF-β1, TNF-α, and S100A11 response in clinical samples from healthy control group (C1-C8) and MASH (M1-M10). Data were adjusted to the total number of EVs measured by NTA. Receiver operator characteristic (ROC) curves on response in control and MASH samples.

As the next step in establishing a diagnostic assay, we proceeded to capture and analyze EVs using SPR. This analytical approach was chosen for the following reasons: 1) it measures binding events directly without additional labeling steps, 2) SPR analysis is performed on Au substrates that are identical in composition (functionalization) to electrodes used in an electrochemical immunoassay, and 3) as an established analytical method, SPR was particularly well-suited for benchmarking our newly developed electrochemical immunosensor. The process of surface functionalization is described in Figure S2 and Figure 2E. It consisted of first forming an alkanethiol layer with terminal carboxyl groups (MUA treatment), then converting the carboxyl groups into amine-reactive intermediates by EDC/NHS treatment and finally incubating surfaces with Ab molecules. Each step in the assembly of this biorecognition layer was monitored using SPR (see Figure S2). Once assembly of all the components was confirmed, we proceeded with EV capture experiments [Figure 2E; (i) EV Capture]. Figure 2F shows SPR sensograms for EV binding on SPR chips presenting anti-ASGR2 Ab surface. Once binding of EVs was confirmed, we proceeded to label EVs with Abs for biomarkers of interest: TNF-α, TGF-β1 and S100A11 [Figure 2E; (ii) Marker Detection]. As shown in Figure 2G, the SPR signals from all three biomarkers were considerably higher in the PA-treated EV group than in the control group. This confirmed our hypothesis that TNF-α, TGF-β1 and S100A11 may be present on EVs originating from injured hepatocytes. We note that Abs for labeling surface markers were introduced sequentially. The sequence of labeling with Abs was determined empirically and was found to work best when labeling less abundant surface markers first (TGF-β1 < TNF-α < S100A11). Table in Figure 2G summarizes SPR responses of EV capture and surface marker detection for different experimental groups described above. These results underscore that the expression of liver injury biomarkers (TGF-β1, TNF-α, and S100A11) was upregulated in hepatic spheroids treated with PA. Additional control experiments (see Figure S3) revealed minimal interactions of captured EVs with rabbit IgG Abs (isotype control).

In summary, our analysis revealed that lipotoxic injury caused upregulation of TNF-α, TGF-β1 and S100A11 at the cellular level and that EVs shed by injured cells were also enriched in these disease biomarkers. With this knowledge in hand, we moved forward to analyzing EVs in patient plasma samples.

### SPR analysis of hepatic EVs in patient plasma samples

While the primary objective of this study was to develop an electrochemical immunosensor for EV-based diagnosis of MASH, it was important to establish an orthogonal benchmarking approach to validate assay performance. SPR represents an excellent validation method for reasons mentioned in the previous section and was used for analyzing a limited cohort of plasma samples collected from patients with MASH and healthy controls.

Figure 2H describes an experimental workflow which involved purifying EVs from plasma using ultracentrifugation followed by SPR analysis on chips functionalized with anti-ASGR2 Abs. Representative NTA results depicting EV concentration and size distribution for healthy control 1 (C1) and MASH 6 (M6) samples are shown in Figure S4. Analysis of all samples (see Table S2) revealed that while the mean particle size for each group was not significantly different, the concentration of EVs in the MASH group was 2.2-fold higher than healthy control groups. This is consistent with previous literature reports (*2*) and our *in vitro* culture results, where injured hepatocytes produced more EVs than uninjured controls. As discussed previously, for SPR assays, EV concentration was adjusted to be the same (2×10^9^ particles/mL) for all samples. Figure 2I shows similar EV binding signals for C1 and M6 samples. In fact, analysis of EV binding for all samples did not reveal significant differences in EV binding depending on the sample group - MASH and healthy controls (see Figure S5 for summary of this experiment). This suggests that ASGR2 expression levels were similar across EVs from all sample groups. It was therefore exciting to observe that, despite capturing similar numbers of EVs per sample group, disease biomarker expression was significantly higher on MASH EVs compared to control EVs (see Figure 2I; (ii) Marker Detection). This led us to conclude that levels of TGF-β1, TNF-α and S100A11 were higher, on per EV basis, for hepatic EVs originating from the injured liver. This observation was consistent with analysis of hepatic EVs from an *in vitro* injury model. The protein expression levels on hepatic EVs are summarized in Figure S5.

As the next step, we wanted to evaluate a scenario where EV concentrations were reflective of plasma concentrations and not adjusted to parity as described in the previous section. We wanted to minimize non-specific binding of plasma proteins when obtaining a benchmarking dataset with SPR, and therefore, isolated EVs from plasma samples prior to analysis. Figure 2J summarizes SPR measurements for TGF-β1, TNF-α and S100A11 on hepatic EVs from patient plasma. These data make it evident that EVs derived from MASH patients exhibited higher expression levels of all three markers. As may be seen from receiver operating characteristic (ROC) curves, the sensitivity and selectivity of the three biomarkers was 100 % for discriminating MASH vs. healthy controls, with an area under the curve (AUC) of 1.0.

### Constructing and characterizing electrochemical immunosensor

Having demonstrated that hepatic EVs from injured cells and a small cohort of patient samples are enriched in injury markers, we proceeded to implement a microtiter-based electrochemical immunoassay for testing a larger cohort of clinical samples. The workflow for EV-based electrochemical detection is illustrated in Figure 3A. Overall, plasma samples from 77 patients were analyzed, including a training cohort of 22 healthy controls and 22 patients with MASH, and a validation cohort consisting of 27 non-MASH liver disease controls and 6 patients with MASH. Patient plasma was diluted 1:100 and introduced into a custom-designed electrochemical microtiter plate containing microwells integrated with Au working electrodes (WEs) for selective capture of hepatic EVs. The design and operating principle of the microtiter plate are shown in Figure 3B. The device comprises 16 working electrodes patterned on a glass substrate, together with on-chip reference and counter electrodes (REs/CEs). A polydimethylsiloxane (PDMS) layer defines individual microwells over each WE and incorporates capillary valves that hydraulically isolate the WEs from the RE/CE. These capillary valves enable independent incubation of WEs with plasma samples and subsequent labeling with immunoprobes while maintaining the REs/CEs in a pristine state (Figure 3B-1). At the measurement stage, the capillary valves are actuated by applying pressure (∼0.7 psi) through the electrolyte inlet, driving the electrolyte into the channel and electrically connecting the WEs with the REs/CEs (Figure 3B-2). This design ensures controlled assay processing and minimizes electrode cross-contamination prior to readout. To selectively isolate hepatic EVs from the heterogeneous population of circulating EVs, WEs were functionalized with Abs against the hepatocyte-specific surface marker ASGR2. Captured EVs were subsequently quantified using a nanoparticle-enabled electrochemical immunoassay. Gold nanoparticles (AuNPs) were functionalized with target-specific Abs and loaded with Pb²⁺ ions to impart redox activity. These AuNP-based immunoprobes (denoted as AuNPs/Abs@Pb²⁺ or immunoprobes throughout the manuscript) were detected using square-wave voltammetry (SWV), enabling sensitive quantification of MASH-associated EV biomarkers. The stepwise assembly of the biorecognition layer on WEs were detailed in Figure S6. Each surface modification step was electrically characterized by electrochemical impedance spectroscopy (EIS), confirming successful electrode functionalization (Figure 3C). This characterization, carried out in 5 mM [Fe(CN)₆]³⁻/⁴⁻ showed that electrode impedance (semicircle diameter) increased as result of self-assembly of 11-mercaptoundecanoic acid (MUA) (black), attachment of Abs (purple) and passivation with casein (green). Incubation with plasma resulted in further increase in the impedance signal (red), suggesting that EV capture hindered interfacial electrode transfer. It is important to note that subsequent labeling with immunoprobes (blue) reduced electrode impedance which was consistent with presence of conductive gold nanoparticles that facilitate electron transfer across the interface. This experiment confirmed steps associated with electrode functionalization, EV capture and surface marker detection. We next evaluated the extent of nonspecific binding on the electrodes after incubating with patient plasma. For this set of experiments (see Figure 3D), some electrodes were functionalized with anti-ASGR2 while other electrodes had non-specific rabbit IgG Abs. Incubation with MASH plasma sample revealed that impedance signals from rabbit IgG electrodes were minimal and comparable to signals before incubation with plasma. Conversely, high impedance signals were observed from anti-ASGR2 electrodes.

**Figure 3.**
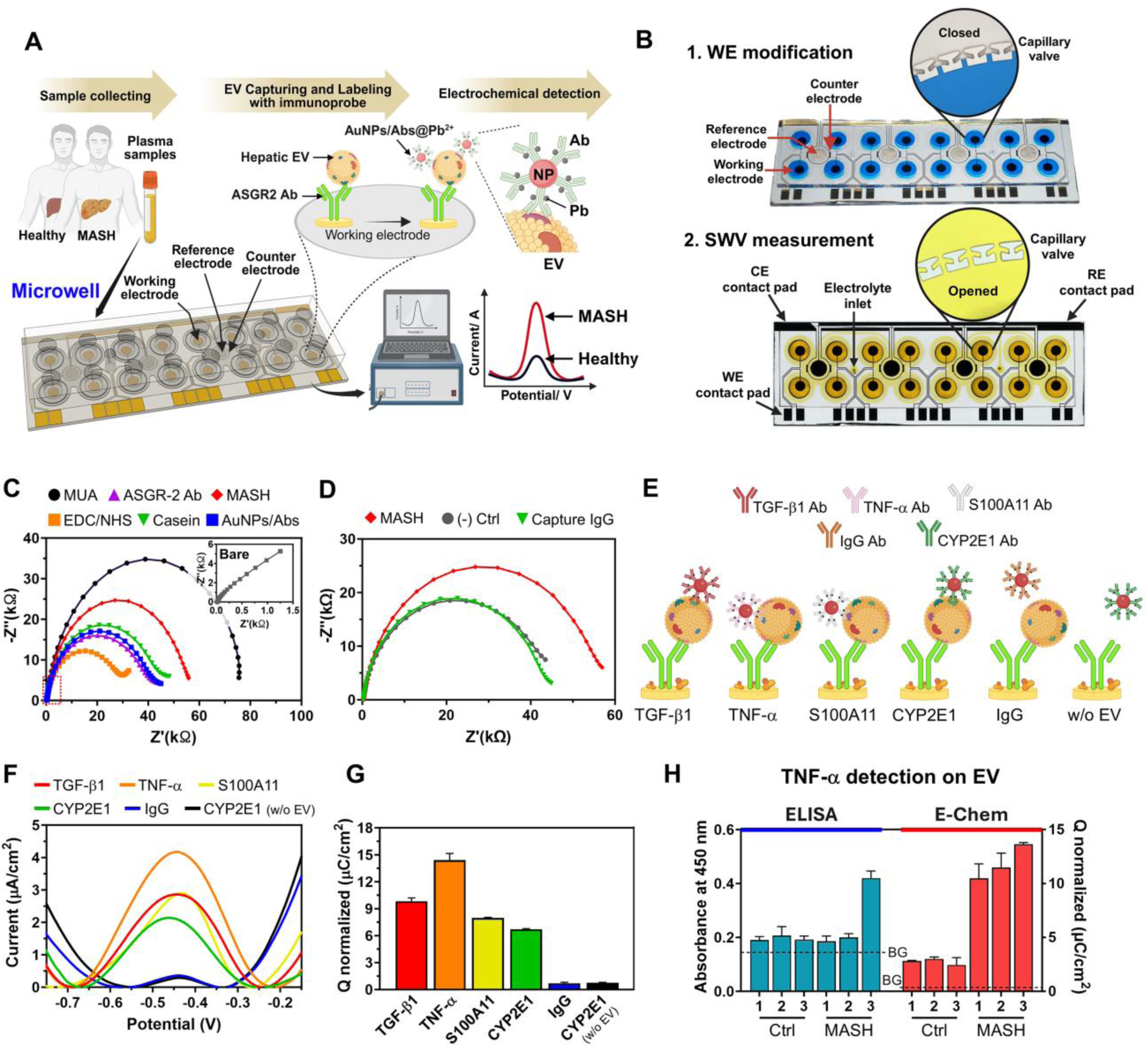
Characterization of the electrochemical immunosensor for hepatic EV capture and multiplexed biomarker analysis. (A) Schematic of the assay workflow. Diluted patient plasma is incubated in microwells containing Au working electrodes functionalized with anti-ASGR2 for selective capture of hepatic EVs. Captured EVs are labeled with AuNP-based immunoprobes carrying redox-active metal ions, and biomarker levels are quantified by square-wave voltammetry (SWV). (B) Design and operation of the electrochemical microtiter plate. A PDMS layer with microwells and capillary valves is integrated with a glass substrate patterned with 16 Au working electrodes (WEs) and on-chip reference (RE) and counter electrodes (CE). Capillary valves enable sequential incubation and washing while isolating RE/CE from WE. Electrochemical measurements are initiated by pressure-driven electrolyte connection. (C) Electrochemical impedance spectroscopy (EIS) characterization of stepwise surface modification: bare Au (grey), Au/MUA (black), Au/MUA/EDC–NHS (orange), Au/MUA/EDC–NHS/anti-ASGR2 (purple), Au/MUA/EDC–NHS/anti-ASGR2/casein (green), Au/MUA/EDC–NHS/anti-ASGR2/casein/EVs (red), and Au/MUA/EDC–NHS/anti-ASGR2/casein/EVs/AuNP–anti-CYP2E1@Pb²⁺ (blue). (D) EIS-based evaluation of hepatic EV capture from plasma of MASH patients, using anti-ASGR2-functionalized electrodes compared to IgG and negative controls. (E) Specificity assessment using immunoprobes targeting CYP2E1, TGF-β1, TNF-α, and S100A11. Negative controls included AuNP–anti-IgG@Pb²⁺ in the presence of EVs and AuNP–anti-CYP2E1@Pb²⁺ in the absence of EVs. (F) Representative SWV curves corresponding to the experimental groups shown in (E). (G) Quantification of total charge (Q), calculated as the area under SWV peaks. Minimal redox signals were observed in negative and isotype control conditions. (H) Comparison of ELISA and electrochemical detection (SWV) for TNF-α quantification on hepatic EVs from control and MASH plasma samples.

Collectively, these EIS results validate the successful construction of the biorecognition layer and demonstrate specific capture of hepatic EVs on the electrode surface.

### Optimizing electrochemical immunosensor in preparation for clinical sample testing

In the next set of experiments we characterized electrochemical immunoassay, focusing on interactions of AuNP-based immunoprobes with electrodes. Human plasma contains a heterogeneous population of EVs along with serum proteins, salts, and other potential interferents that may influence electrochemical readout. To ensure robust performance, we systematically optimized key assay parameters, including surface blocking, plasma dilution, and matrix effects. For assay validation, AuNP-based immunoprobes were functionalized with Abs targeting the hepatic marker CYP2E1 (used for assay optimization) and the MASH-associated biomarkers TGF-β1, TNF-α, and S100A11. Rabbit IgG-functionalized immunoprobes served as isotype controls (Figure 3E), while AuNPs@CYP2E1@Pb^2+^ applied in the absence of plasma served as negative controls. (Figure 3E). Representative square-wave voltammetry (SWV) curves (Figure 3F) and corresponding total charge quantification (area under the SWV curve; Figure 3G) showed significantly higher electrochemical responses for CYP2E1 and MASH-associated biomarkers compared to control conditions, confirming assay specificity and minimized non-specific binding of immunoprobes following the washing protocol.

Electrode surface blocking was critical to minimizing non-specific adsorption and improving the sample-to-blank ratio. Casein was selected as the blocking reagent over bovine serum albumin (BSA) due to its superior blocking performance (Figure S7). Optimization of plasma dilution revealed that a 1:100 dilution in SuperBlock buffer provided a robust and reproducible signal while minimizing matrix effects arising from plasma proteins and ionic species (Figure S8).

To further assess matrix effects, we compared electrochemical responses from purified EVs, EV-depleted plasma, and diluted whole plasma (Figure S9). Purified EVs produced higher electrochemical responses than whole plasma, consistent with reduced protein interference at the electrode surface. Importantly, diluted plasma generated significantly higher signals than EV-depleted plasma, confirming that electrochemical signals originate from EV-associated biomarkers. EV-depleted plasma showed signals comparable to IgG-functionalized controls, further supporting assay specificity. We further compared electrochemical responses between purified EVs and diluted plasma for TGF-β1, TNF-α, and S100A11 (Figure S10). Although signal intensity was reduced in diluted plasma relative to purified EVs, the relative expression trends were preserved, demonstrating reliable detection regardless of plasma sample complexity. Collectively, these results demonstrate sensitive and specific detection of immunoprobe binding to EVs and position us for clinical sample testing using electrochemical immunoassay.,

We next asked how our electrochemical immunoassay compares to more traditional colorimetric enzyme linked immunosorbent assays (ELISAs). Because no commercial ELISA for capturing hepatic EVs and analyzing desired EV surface markers existed, we developed our own colorimetric ELISA assay following assay design described by Lee et al. (*34*). Design of this custom colorimetric ELISA is described in Figure S11. It involved (1) immobilization of anti-ASGR2 on the 96-well plate, (2) incubation with patient plasma to capture hepatic EVs, (3) incubation with detection antibody (biotinylated anti-TNF-α), (4) followed by incubation with streptavidin-HRP and colorimetric substrate. Given limited availability of patient plasma, and larger volumes required for standard 96-well plate compared to our electrochemical microtiter plate (100 µL vs 50 µL), we focused on only one biomarker. TNF-α was chosen because it produced strongest signals in hepatocyte-derived EVs when tested by SPR and would be most likely to be detected with ELISA. We selected three MASH samples with TNF-α in the high (F3), medium (F2) and low range (F0-1) as well as three control plasma samples. As may be appreciated from Figure 3H, signals from all control samples and two MASH samples were similar to the background (diluent solution without plasma). Only the sample with high TNF-α signal could be distinguished using ELISA. Conversely, with electrochemical immunoassay, all six samples were significantly higher than the background, and MASH samples were significantly different than controls. These results are in alignment with work of Lee et al. who also reported electrochemical detection of EV surface markers to be superior to optical detection with ELISA (*34*). The superior sensitivity of the electrochemical immunosensor may be attributed to a combination of factors including high sensitivity of nanoparticle-based immunoprobes and low background signal due to high quality functionalization of the Au electrode surface.

### Testing patient plasma using electrochemical immunosensor

Given the robust analytical performance of the electrochemical immunosensor discussed above, we next evaluated its diagnostic utility in a training cohort comprising 22 patients with MASH and 22 healthy controls (n = 44). Figure 4A summarizes EV expression profiles of all MASH-associated biomarkers across this cohort. As shown in Figures 4B–4D, circulating hepatic EVs from MASH patients exhibited significantly elevated expression of the three MASH-associated biomarkers—TGF-β1, TNF-α, and S100A11—relative to healthy controls (p < 0.0001 for all comparisons). Receiver operating characteristic (ROC) analysis demonstrated excellent diagnostic performance for each individual marker (Figure S12), with an area under the curve (AUC) of 1.0, corresponding to 100% sensitivity (positive predictive agreement) and 100% specificity (negative predictive agreement) in distinguishing MASH from healthy controls within the training cohort. To validate performance using orthogonal analytical method, electrochemical immunoassay was benchmarked against SPR using 18 plasma samples (MASH and healthy controls). Strong correlations were observed across all biomarkers, with Pearson correlation coefficients of 0.86 (TGF-β1), 0.91 (TNF-α), and 0.90 (S100A11) (Figure S13), confirming the robustness and analytical reliability of the electrochemical platform.

**Figure 4.**
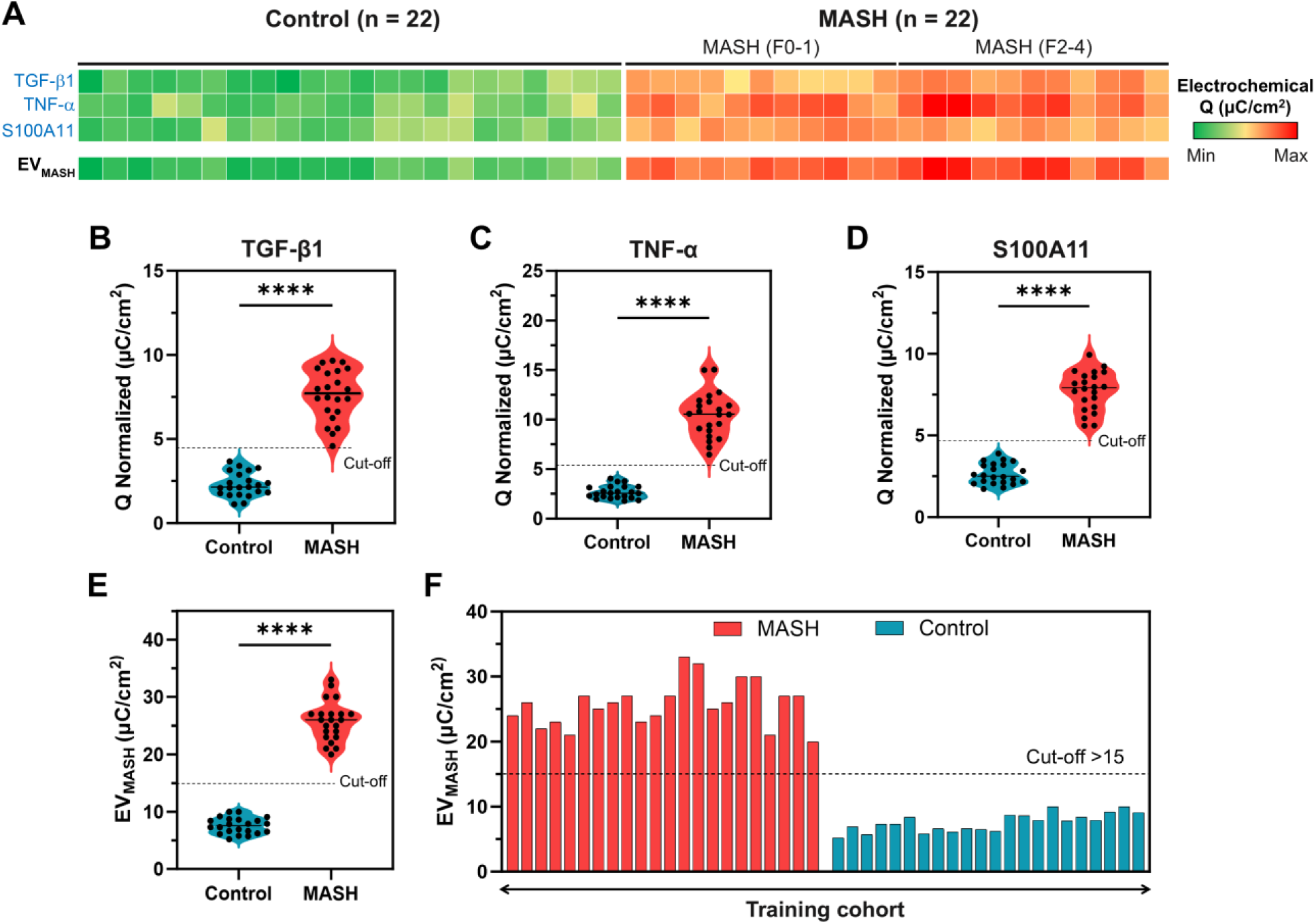
Electrochemical immunoassay of hepatic EVs is used to differentiate MASH from healthy controls. (A) Heatmap of electrochemical measurements from plasma samples of 22 healthy controls and 22 patients with MASH. Expression levels of TGF-β1, TNF-α, and S100A11 on hepatic EVs were quantified by square-wave voltammetry (SWV). The composite diagnostic score (EV_MASH_) was calculated as the sum of the three biomarker signals. (B–D) Quantification of individual biomarkers demonstrating significantly elevated levels in MASH patients compared with controls (p < 0.0001 for all markers; unpaired two-tailed t-test). (E) Composite EV_MASH_ score showing clear separation between MASH and control groups (p < 0.0001; unpaired two-tailed t-test). (F) Diagnostic classification in the training cohort based on ROC-derived cut-off value (EV_MASH_ = 15), yielding 100% sensitivity, 100% specificity, and 100% overall accuracy.

Because MASH pathogenesis involves the combined processes of fibrosis, inflammation, and steatosis, we reasoned that integrating these biologically relevant biomarkers into a composite score may provide a more comprehensive representation of disease state. We therefore generated a multi-marker index (EV_MASH_) by combining expression levels of TGF-β1, TNF-α, and S100A11 and performed ROC analysis (Figure 4E; Figure S14). An optimized cut-off value of 15 was determined from ROC analysis (maximizing the Youden index) of the EV_MASH_ score in the training cohort, the model achieved 100% sensitivity, specificity, and overall accuracy in distinguishing MASH from healthy controls (Figure 4F; Table S3). While individual biomarkers already demonstrated strong performance in this cohort, the composite score provides an integrated framework that captures multiple features of disease pathogenesis and supports subsequent analysis in more clinically heterogeneous populations.

We next evaluated electrochemical immunoassay using an independent validation cohort (n = 33), including 6 patients with MASH and 27 patients with non-MASH liver conditions (obese normal (ON), simple steatosis (SS), primary sclerosing cholangitis (PSC), and primary biliary cholangitis (PBC)). In this study, histological assessments of steatosis, inflammation, and fibrosis stage were obtained from liver biopsy specimens and scored by an expert hepatopathologist for all MASH, ON, and SS patients (Tables S4–S6). Cholestatic liver disease patients were similarly diagnosed with liver biopsy and staged according to the Ludwig classification system for PBC and PSC, with detailed histopathological scores summarized in Table S7. As shown in Figure 5A, MASH patients consistently showed elevated levels of all three biomarkers relative to other disease groups. Single-marker analyses (Figure 5B–5D) revealed statistically significant separation between MASH and most non-MASH conditions, with the exception of S100A11 in simple steatosis, likely reflecting the shared steatotic component between these two groups. ROC analysis revealed variable performance among individual biomarkers in this more heterogeneous cohort (Figure S15). The AUC for TGF-β1 decreased to 0.98, with 92.6% sensitivity and 100% specificity. S100A11 showed reduced performance, with an AUC of 0.87 and corresponding sensitivity and specificity of 74% and 83%, respectively. In contrast, TNF-α maintained strong performance, with an AUC of 1.0 and 100% sensitivity and specificity. Importantly, the composite EV_MASH_ score achieved near-perfect diagnostic performance (AUC ≈ 1.0), with 100% sensitivity and specificity, outperforming individual biomarkers and highlighting the advantage of the multi-marker strategy for MASH diagnosis, particularly in the setting of increased clinical heterogeneity (Figure 5E, Figure S16). When the cut-off value determined in the training cohort was applied to the validation cohort (Figure 5F), EV_MASH_ maintained strong diagnostic performance (100% sensitivity, 88.9% specificity, 90.9% overall accuracy). These findings indicate that while single biomarkers may partially overlap across disease states, integrated electrochemical profiling of multiple biomarkers enables accurate classification of MASH within a clinically diverse population, including simple steatosis and cholestatic liver diseases. To further visualize group separation, we applied t-distributed stochastic neighbor embedding (t-SNE) to the combined electrochemical signatures (Figure 5G). Unlike the EV_MASH_ score, t-SNE offers an unsupervised, low-dimensional projection of multiple markers, identifying the inherent similarity without prior group labeling. In this analysis, MASH samples formed a distinct and compact cluster that was clearly segregated from non-MASH groups, whereas control and other liver disease subtypes exhibited a partial overlap. To quantitatively assess cluster quality of t-SNE clusters, silhouette scores were calculated for each disease group, and the results are summarized in Table S8. The MASH cluster exhibited the highest silhouette score (+0.76 ± 0.05), demonstrating the MASH data has strong intra-group similarity and clear separation from other liver disease types. Considering the non-MASH groups, SS showed the second-highest silhouette score (+0.58 ± 0.15), suggesting a relatively distinct electrochemical signature despite sharing steatotic features with MASH. In contrast, PBC and PSC exhibited overlap with healthy controls and other disease groups, consistent with the distinct pathophysiology of cholestatic liver diseases relative to MASH.

**Figure 5.**
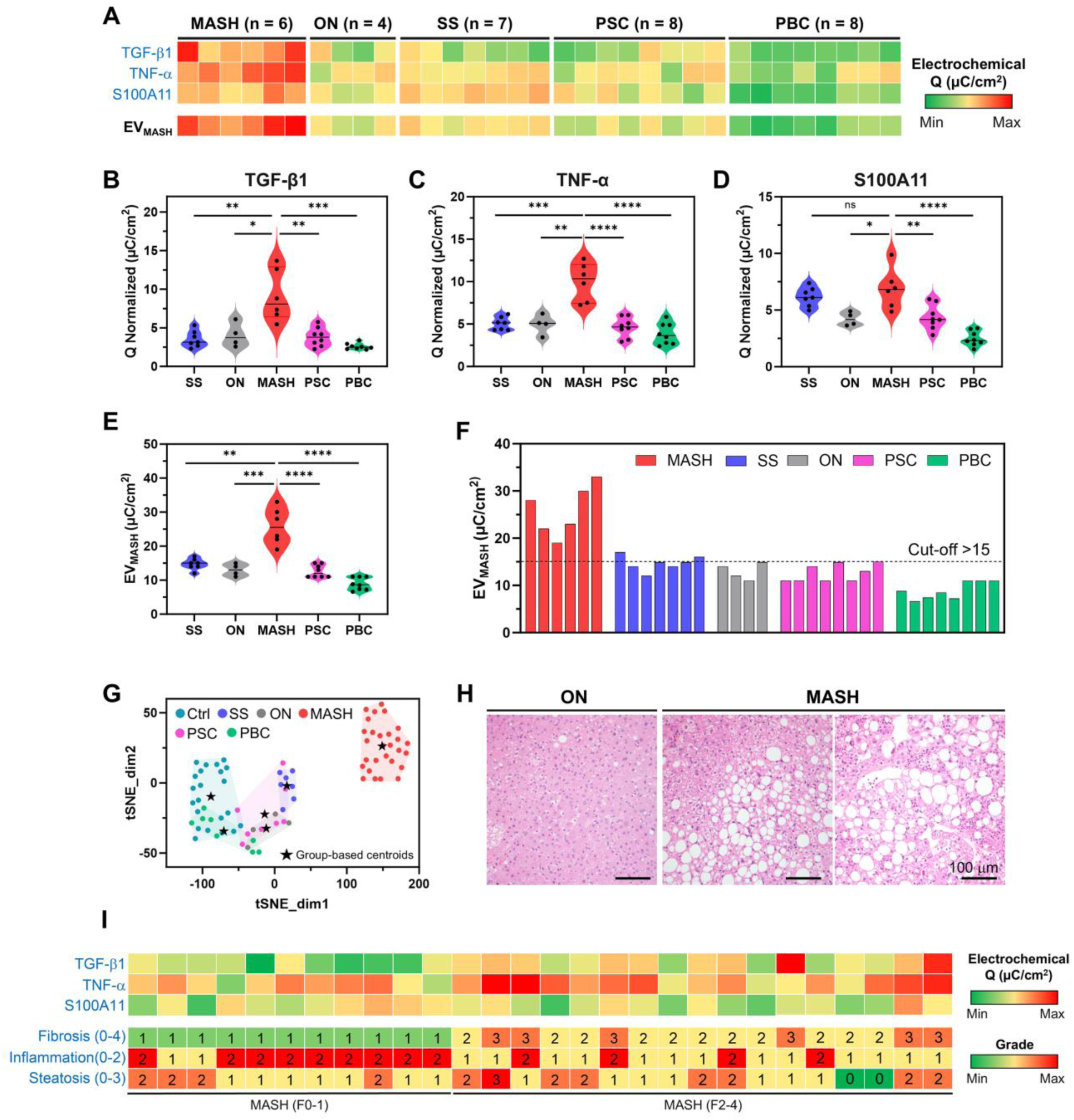
Electrochemical immunoassay of hepatic EVs is used to differentiate MASH from cholestatic liver diseases and other patient populations. (A) Heatmap of electrochemical measurements from the independent validation cohort comprising 6 patients with MASH and 27 patients with non-MASH liver diseases (obese controls [ON], n = 4; simple steatosis [SS], n = 7; primary sclerosing cholangitis [PSC], n = 8; primary biliary cholangitis [PBC], n = 8). Expression levels of TGF-β1, TNF-α, and S100A11 on hepatic EVs were quantified by square-wave voltammetry (SWV). The composite score (EV_MASH_) was calculated using the same formula derived from the training cohort. (B–D) Quantification of individual biomarkers demonstrating significantly elevated levels in MASH compared to non-MASH groups (p < 0.05 to p < 0.0001; unpaired two-tailed t-test), except for S100A11 in comparison with SS. (E) Composite EV_MASH_ score showing significant separation between MASH and non-MASH groups (p < 0.01 to p < 0.0001; unpaired two-tailed t-test). (F) Diagnostic classification using the ROC-derived cut-off value (EV_MASH_ = 15) established in the training cohort, yielding >90% overall accuracy in the validation cohort. (G) t-distributed stochastic neighbor embedding (t-SNE) analysis of electrochemical signals demonstrating clear clustering trends distinguishing MASH samples from non-MASH groups (n = 77, features = 3, perplexity = 10, learning rate = 200). The stars denote the centroids of each disease group. (H) Representative H&E-stained liver sections from obese control (ON) and MASH patients. (I) Correlation between electrochemical EV biomarker levels (upper panel) and corresponding histological grades of fibrosis, inflammation, and steatosis (lower panel) in MASH patients.

To investigate whether our electrochemical immunoassay could capture disease severity, MASH samples were further characterized according to fibrosis stage and analyzed as separate clusters (Figure S17, Table S9). We calculated the silhouette scores and found that the F3 MASH stages formed a distinct cluster with a mean silhouette score of +0.47 ± 0.12, whereas F1 and F2 MASH stages display significant overlap, with mean silhouette scores of +0.16 ± 0.31 and −0.04 ± 0.28, respectively. These findings suggest that analysis of hepatic EVs may reveal differences associated with stage of fibrosis within MASH. We acknowledge that testing a larger patient cohort will be necessary to confirm this observation. Collectively, the t-SNE analysis provides orthogonal, visualization-based support for the discriminatory power of multiplexed electrochemical EV profiling.

As a final step in evaluating the clinical utility of our liquid biopsy approach, we compared plasma-based electrochemical immunosensor results with the current gold standard diagnosis established by liver biopsy and histopathological assessment. Representative H&E-stained liver sections from ON and patients with MASH are shown in Figure 5H. As expected, liver tissue from MASH patients showed the defining histopathological hallmarks of the disease, including macrovesicular steatosis, hepatocyte ballooning, lobular inflammation, and steatohepatitis, features that collectively inform fibrosis staging and inflammatory grading. We next correlated EV profiling of TGF-β1, TNF-α and S100A11 with histological staging parameters from biopsy evaluation (Figure 5I). Increased TGF-β1 levels on circulating hepatic EVs were associated with advanced fibrosis (stages 2–4), whereas lower expression corresponded to mild or no fibrosis (stages 0–1). Similarly, TNF-α levels on hepatic EVs positively correlated with lobular inflammation scores, and S100A11 expression aligned with the degree of steatosis. These concordant relationships between EV profiling using electrochemical measurements and tissue-level pathology substantiate the biological relevance of the selected biomarkers and support their utility in detecting underlying disease severity.

## DISCUSSION

In this study, we developed and clinically validated an electrochemical immunosensor for diagnosis of metabolic dysfunction–associated steatohepatitis (MASH). Our work integrates biological discovery with multiple innovations. We established an in vitro lipotoxicity model using primary human hepatocytes to generate hepatic EVs enriched in disease-associated biomarkers. Injured hepatocytes released EVs with significantly upregulated expression of TGF-β1, TNF-α, and S100A11, supporting their biological relevance to fibrosis, inflammation, and steatosis. SPR was used to optimize EV capture chemistry and independently validate EV surface marker expression. Building on this foundation, we implemented a custom microtiter plate as an electrochemical immunosensor for detecting hepatic EVs in patient plasma. This approach enabled (i) capture of hepatic EVs from whole plasma without prior purification, (ii) sensitive detection of MASH-associated biomarkers on hepatic EVs via AuNP-enabled metal ion signal amplification, and (iii) analysis of multiple biomarkers within a compact formfactor device suitable for clinical translation. Compared with ELISA, the electrochemical platform demonstrated substantially enhanced sensitivity and dynamic range. 77 patient plasma samples were tested to assess diagnostic performance of the electrochemical immunoassay. In the training cohort, individual biomarkers and the composite EV_MASH_ score achieved 100% sensitivity, specificity and accuracy for differentiating MASH from healthy controls. When the model was applied to an independent validation cohort containing MASH and diverse non-MASH liver diseases, including simple steatosis and cholestatic liver conditions, the electrochemical immunoassay maintained strong diagnostic performance, achieving overall 90.9% accuracy based on the EV_MASH_ score. Applying t-SNE to the electrochemical data allowed MASH to be clearly differentiated from non-MASH diseases. Furthermore, EV biomarker levels correlated with histological staging parameters derived from liver biopsy, supporting the biological and clinical relevance of the platform Although liver biopsy remains the clinical gold standard for MASH diagnosis and staging, histological grading is inherently semi-quantitative and subject to sampling bias and inter-observer variability. Moreover, liver biopsy is invasive, costly, and not routinely performed in all patients. The minimally invasive liquid biopsy approach described here requires only a small volume of plasma and provides a quantitative measure of liver disease. While non-invasive imaging methods such as magnetic resonance elastography are emerging, they remain costly, infrastructure-dependent, and inaccessible in many settings. In contrast, our electrochemical platform is compact, scalable, and potentially deployable in resource-limited environments, positioning it as a promising tool for population-level screening, repeated longitudinal disease monitoring, and therapeutic response assessment. Additionally, our platform is pathophysiologically informed and not influenced by BMI, age, and aspartate aminotransferase levels, which are common components of noninvasive tests for identifying at-risk MASH. We acknowledge that larger, multi-center studies and longitudinal validation will be necessary to further define clinical utility, including evaluation for treatment monitoring and risk stratification and testing in younger patients. Nevertheless, given that approximately 30% of the global population is affected by MASLD and ∼5% by MASH—most of whom remain undiagnosed—the need for accessible, scalable diagnostics is urgent. With the recent approval of therapeutic agents for MASH patients with fibrosis stage 2 and 3, and multiple agents advancing through late-stage clinical trials, there is a critical need for reliable tools to identify eligible patients and monitor therapeutic response. Our electrochemical immunosensor platform was developed to address this critical need by leveraging key aspects of disease pathogenesis to enable a practical, scalable, liquid biopsy–based solution for MASH diagnosis.

## MATERIALS AND METHODS

### Study design

This study aimed to develop a minimally invasive liquid biopsy platform for MASH diagnosis by targeting disease-specific biomarkers on circulating hepatic EVs. We first established an in vitro lipotoxic injury model in hepatocytes to confirm that hepatic EVs carry biomarkers associated with steatosis (S100A11), inflammation (TNF-α), and fibrosis (TGF-β1). The presence of these biomarkers was subsequently validated using an electrochemical immunoassay using patient plasma. In this study, we developed a custom electrochemical microtiter plate integrating working electrodes, microfluidic channels, and capillary valves to enable direct capture of hepatic EVs from whole plasma and multiplexed detection of EV surface markers. The analytical performance of our platform was compared against conventional ELISA and cross-validated SPR analysis using EVs isolated from same plasma. Finally, clinical performance of the developed electrochemical immunosensor was evaluated using 77 human plasma samples, comprising a training cohort (n = 44; 22 MASH patients and 22 healthy controls) and an independent validation cohort (n = 33; 6 MASH and 27 non-MASH, including simple steatosis and cholestatic liver diseases). The electrochemical immunosensor achieved 100% sensitivity, specificity, and accuracy in the training cohort, and maintained >90% diagnostic accuracy in the independent validation cohort despite increased clinical heterogeneity. All human plasma samples were collected under Mayo Clinic Institutional Review Board approval with informed consent from all participants.

### *In vitro* lipid liver injury model

Primary human hepatocytes were cultured in T75 flasks in EV-free FBS containing DMEM media described in the previous paragraph. Then, PA in isopropyl alcohol (IPA) was added into EV-free media to achieve a final concentration of to 0.4 mM (*48*) The cells were cultured for 72 h with daily changes of media (with or without PA depending on the experimental group).

### Isolation and characterization of EVs from cell-conditioned media

#### Isolation of EVs

In this study, small EVs were separated from large EVs using differential ultracentrifugation. Small EVs were then separated from soluble proteins and protein aggregates by sedimentation in a density gradient. Briefly, conditioned media was first centrifuged at 500 g for 10 min to remove cellular debris. The supernatant was transferred to a fresh microcentrifuge tube and centrifuged at 20,000 g for 30 min. The resulting supernatant was carefully layered onto a sucrose-PBS cushion and subjected to ultracentrifugation at 100,000 g for 90 min. Finally, small EVs were collected from the interface fractions, which corresponded to sucrose concentrations ranging from 6% to 60%.

#### Characterization of EVs

Nanoparticle Tracking Analysis (NTA) was performed using a NanoSight NS300 (Malvern Panalytical) to assess the concentration and particle size of the isolated EVs. EV morphology was characterized by scanning electron microscopy (SEM) using a Hitachi S-4700 cold field emission SEM (Hitachi High Technologies America, Inc.). For Western blot (WB) analysis, EVs were mixed with an equal volume (1:1, v:v) of 2× Laemmli buffer (containing β-mercaptoethanol) and heated at 95°C for 10 min. The samples were then run on a 12.5% SDS-PAGE gel, followed by transfer to a 0.22 µm PVDF membrane using Tris-Glycine buffer. The membrane was blocked with 5% milk in TBST buffer and incubated overnight with a primary antibody, diluted 1:1000 in 5% BSA, at 4°C. After several washes with TBST, the membrane was incubated with a secondary antibody, diluted 1:3000, at room temperature for 2 h. After additional TBST washes, the membrane was developed using ECL reagent for signal detection.

### Human plasma samples

The human studies were conducted following approval by the Mayo Clinic Institutional Review Board. All participants provided written informed consent to participate in medical research. Biobanked patient samples from MASH and obese normal were from previously described cohorts (*49–51*). Health control plasmas were collected from a Mayo Clinic Biobank. Plasma samples from cholestatic patients were obtained from the PSC Scientific Community Resource, which includes individuals with PSC, PBC, and healthy controls (*52*). The diagnosis of PSC or PBC was established by expert pathologist review of a clinically indicated liver biopsy per the Ludwig staging system (*53*). In this group, fibrosis ranged from 0-4.

### EV isolation from patient plasma samples

50 µL of plasma was diluted 10 times with 1× PBS and precleaned to remove debris by centrifugation at 2,000 g for 30 min at 4°C. The supernatant was collected and centrifuged again at 12,000 g for 45 min at 4°C to obtain large EV. After large EVs were removed, the solution was processed by ultracentrifugation at 110,000 g for 2 h at 4°C. The pellet was collected and dispersed in 100 µL 1× PBS. The small EV was aliquoted based on the NTA results and stored at −80°C.

### SPR analysis

SPR chips (Biosensing Instrument) were cleaned with IPA for 20 min under sonication. Then, a chip was treated with oxygen plasma for 3 min at 150 mW (YES-G500, Yield Engineering Systems, Freemont, CA, USA). A chip was then immersed in 10 mM MUA solution in pure ethanol overnight to create a well-ordered alkanethiol monolayer with carboxylic acid end functional groups. A chip functionalized with MUA was washed with ethanol 3 times, dried with N2 gas and placed into an SPR instrument (BI-2500; Biosensing Instruments). An SPR chip with MUA layer was further functionalized by first injecting EDC/NHS solution for surface activation, then introducing 20 µg/mL of anti-ASGR2 in 50 mM MES buffer and finally flow in 1% casein in 1× PBS for blocking surface (w/v). Then, 1×10^9^ particles/mL of cell-derived EV or 2×10^9^ particles/mL of plasma EV in 1× PBS with 6% sucrose were injected for 10 min. Finally, detection Abs (anti-TGF-β1, anti-TNF-α and anti-S100A11) at 10 µg/mL in 1x PBS were infused into flow cell to characterize expression of surface markers on captured EVs. To assess the extent of non-specific interactions between EV and labeling Abs, isotype control (rabbit-anti-IgG) antibody solution was injected as the final step. The running buffers for SPR experiments were DI water or 1× PBS, the flow rate for all SPR steps of experiments was 20 µL/min, except EV loading which was performed at 10 µL/min.

### Fabrication of the electrochemical microtiter plate

The microtiter plate was designed using CAD software (AutoCAD 2020, Autodesk Inc.) and fabricated using standard photolithography and metal etching techniques. The microtiter plate consisted of two layers: gold electrodes on glass and PDMS layers containing wells, channels, and capillary valves, as described previously (*15*). The gold electrodes were fabricated by a standard metal sputtering and chemical etching processes. The Ag/AgCl reference electrode was carefully applied to the gold surface and cured at 120°C for 20 min according to the manufacturer’s instructions. The plate featured 16 circular working electrodes (2.5 mm in diameter), 4 reference electrodes, and 4 counter electrodes. All electrodes were connected via 20 μm connection lines to contact pads located at the edges of the glass substrate.

The PDMS microwell layer was fabricated using a wafer master mold. Briefly, PDMS prepolymers (10:1 elastomer/curing agent ratio) were poured onto the mold to a height of approximately 2 mm, degassed in a vacuum chamber, and baked at 80°C for 60 min. Afterward, the PDMS layer was peeled off from the mold, and holes for wells and electrolyte inlets were punched. The fabrication process was completed by placing the PDMS layer onto the electrode array to create 16 independent electrochemical cells, each with a maximum filling volume of 50 μL.

### Functionalization of electrodes and preparation of AuNPs/Ab@Pb^2+^ immunoprobes

The Au working electrodes were functionalized with anti-ASGR2 Abs for EV capture using the following procedure. The Au electrodes were immersed in 10 mM MUA in ethanol for 12 h to allow self-assembly of the alkanethiol. Afterward, the electrodes were washed with ethanol and DI water, then dried with nitrogen. Next, the PDMS layer was aligned and placed onto the glass substrate with the electrode array. The Au electrodes in each well were then treated with a 1:1 ratio of 200 mM EDC and 100 mM NHS in MES buffer (0.1 M, pH 5) for 30 min to create amine-reactive groups. After the microtiter wells were rinsed with DI water, the electrodes were incubated with 25 μg/mL of anti-ASGR2 for 90 min. Following this, the wells were washed with 1× PBS to remove excess Abs. To prevent nonspecific binding, the working electrodes in the wells were blocked with 1% casein for 1 h. After another washing step with 1× PBS, the functionalized microtiter plate was stored at 4°C until further use.

We have previously described a protocol for conjugating AuNPs/Abs@Pb²⁺ (*54*). In this study, we followed this protocol with minor changes to synthesize immunoprobes targeting several EV surface antigens including AuNPs/anti-CYP2E1@Pb²⁺, AuNPs/anti-TNF-α@Pb²⁺, AuNPs/anti-TGF-β1@Pb²⁺, AuNPs/anti-S100A11@Pb²⁺, and AuNPs/anti-IgG@Pb²⁺, which served as a negative (isotype) control.

Briefly, NHS-activated AuNPs (6.54×10¹¹ NPs/mL) were dispersed in 90 μL of buffer containing 40 μg of antibody (Ab) solution and incubated for 4 h at room temperature. Afterward, 10 μL of quencher solution was added to eliminate any remaining unreacted NHS on the AuNPs. To remove unbound Abs and other reagents, the mixture was centrifuged at 7,000 g and 4°C for 20 min and washed three times with HEPES buffer containing 0.05% Tween 20 (0.02 M, pH 7.0), then stored at 4°C until use.

Next, the AuNPs/Ab conjugates were dispersed in 1 mL of HEPES buffer (0.02 M, pH 7.0) with 0.05% Tween 20, followed by the addition of 20 μL of 10 mM Pb(NO₃)₂ aqueous solution. The mixture was stirred overnight, allowing the Pb²⁺ ions to complex with the amine groups of the Abs. Finally, the immunoprobes (AuNPs/Abs@Pb²⁺) were collected by centrifugation, washed thoroughly with HEPES buffer containing 0.05% Tween 20, and redispersed in 1 mL of HEPES buffer (0.02 M, pH 7.0). The immunoprobes were then ready for use.

### Testing patient plasma using electrochemical immunosensor

The plasma samples were diluted 100x in SuperBlock buffer and directly added to the anti-ASGR2-modified electrodes on the microtiter plate. After incubating for 90 min, each well was washed with 1× PBS, followed by the addition of 50 μL of target-specific immunoprobes for a 60-min incubation. Afterward, the wells were washed three times with 1× PBS and DI water, and the samples were analyzed using square wave voltammetry (SWV). The total time for the assay was 150 min (2.5 h). All electrochemical EV measurements were conducted in the same condition as described above.

Electrochemical detection was carried out by connecting the working (Au), counter (Au), and reference (Ag/AgCl) electrodes to a potentiostat (PalmSens). SWVs were recorded in acetic acid/sodium acetate buffer (HAc/NaAc; 0.2 M, pH 4.5) within the −0.8 to 0.0 V range (versus Ag/AgCl), with a 25mV amplitude and a 15 Hz frequency. Each well was filled with 50 μL of electrolyte for electrochemical detection. The electrodes were connected to the potentiostat using a custom-designed clip to the connection pad. Electrochemical signals were recorded from three different working electrodes and are reported as the mean ± SD. To quantify the electrochemical signals, SWV curves were integrated to determine the total charge (Q) under the redox curve. The Q values were then normalized using the formula: normalized Q, Q̅ = Q__EVs_ – Q _isotype control_ Abs).

### TNF-α detection on plasma EV by sandwich ELISA

The ELISA method was designed to detect TNF-α on EV and compare sensitivity of marker detection with our electrochemical analysis. First, 5 µg/mL of anti-ASGR2 Ab in 1× PBS was added in each well of an ELISA plate for overnight at 4°C to immobilize capture Abs. Next day, the wells were rinsed with 1× PBS 3 times. Afterwards, 1% BSA in PBS was added to each well for blocking surface for 1 h at room temperature. Each plasma sample from patient was diluted 100 times with superblock and added 100 µL to each well for 2 hours under shaking. After incubation of plasma samples, 200 µg/mL of biotinylated-TNF-α antibody (R&D systems) was added to each well for 1 h. Then, the Streptavidin-HRP solution (R&D systems) and TMB solution (Sigma-Aldrich) were incubated for 1 h and 20 min, respectively. Once the solution turned blue color, 2 M of sulfuric acid was treated to stop the reaction, and the absorbance was measured by plate reader at a 450 nm wavelength.

### Statistical analysis

All experiments were performed with a minimum of three replicates. For SPR analysis, the responses from three different channels were average. To assess the significance of differences between two groups, unpaired two-tailed t-test was applied. For comparisons involving more than three groups, one-way ANOVA followed by Tukey’s post hoc test was conducted. Statistical significance between groups was denoted as follows: ns (non-significant) for p > 0.05, * for p < 0.05, ** for p < 0.01, *** for p < 0.001, and **** for p < 0.0001. All statistical analyses and graph configurations were performed using GraphPad software.

### T-SNE analysis

Dimensionality reduction was performed using t-distributed stochastic neighbor embedding (t-SNE) on a dataset of 77 electrochemical readouts to enable visualization of underlying patterns. Prior to analysis, all features were standardized using StandardScaler from scikit-learn (v1.6.1) in Python to ensure consistent scaling. The t-SNE model was implemented in Spyder 6.0.7 with a perplexity of 10 and a learning rate of 200 to generate two-dimensional projections for cluster visualization.

## Data Availability

All data produced in the present work are contained in the manuscript

## Acknowledgements

Thi Thanh-Qui Nguyen and Daheui Choi contributed equally to this work.

## Author contributions

T. T.-Q. Nguyen and D. Choi performed experiments and manuscript preparation. S. Lee, J. M. de Hoyos-Vega, P. V. Daniel and A. S. Mauer assisted in characterization and experiment measurement. G. Stybayeva supervised the experiments. M. J. Eller and A. Giron performed t-SNE analysis and worked on manuscript preparation. R. P. Graham provided patient histology data. A. M. Allen and K. N. Lazaridis provided patient plasma and clinical data and worked on manuscript preparation. H. Malhi and A. Revzin conceived and supervised the project. All authors reviewed and approved the manuscript.

## Data and materials availability

All data are available in the main text or the supplementary materials.

## References and Notes

1. S. Wang, S. L. Friedman, Found in translation—Fibrosis in metabolic dysfunction– associated steatohepatitis (MASH). Science translational medicine 15, eadi0759 (2023).

2. Y. Nakao, P. Amrollahi, G. Parthasarathy, A. S. Mauer, T. S. Sehrawat, P. Vanderboom, K. S. Nair, K. Nakao, A. M. Allen, T. Y. Hu, Circulating extracellular vesicles are a biomarker for NAFLD resolution and response to weight loss surgery. Nanomedicine: Nanotechnology, Biology and Medicine 36, 102430 (2021).

3. L. A. van Kleef, J. Pustjens, M. Savas, I. Ayada, P. Li, Q. Pan, E. F. van Rossum, H. L. Janssen, W. P. Brouwer, MASLD, At-Risk MASH and increased liver stiffness are associated with young adulthood obesity without residual risk after losing obesity. Liver International 45, e16169 (2025).

4. L. Henry, J. Paik, Z. M. Younossi, the epidemiologic burden of non-alcoholic fatty liver disease across the world. Alimentary pharmacology & therapeutics 56, 942–956 (2022).

5. J. Cathcart, R. Barrett, J. S. Bowness, A. Mukhopadhya, R. Lynch, J. F. Dillon, Accuracy of Non-Invasive Imaging Techniques for the Diagnosis of MASH in Patients With MASLD: A Systematic Review. Liver International 45, e16127 (2025).

6. V. Lekakis, G. V. Papatheodoridis, Natural history of metabolic dysfunction-associated steatotic liver disease. European Journal of Internal Medicine 122, 3–10 (2024).

7. X. Wang, L. Zhang, B. Dong, Molecular mechanisms in MASLD/MASH-related HCC. Hepatology 82, 1303–1324 (2025).

8. S. K. Bansal, M. B. Bansal, Pathogenesis of MASLD and MASH–role of insulin resistance and lipotoxicity. Alimentary Pharmacology & Therapeutics 59, S10–S22 (2024).

9. R. Itier, M. Guillaume, J.-E. Ricci, F. Roubille, N. Delarche, F. Picard, M. Galinier, J. Roncalli, Non-alcoholic fatty liver disease and heart failure with preserved ejection fraction: from pathophysiology to practical issues. ESC heart failure 8, 789–798 (2021).

10. A. Leszczynska, C. Stoess, H. Sung, D. Povero, A. Eguchi, A. Feldstein, Extracellular vesicles as therapeutic and diagnostic tools for chronic liver diseases. Biomedicines 11, 2808 (2023).

11. Y. Sumida, A. Nakajima, Y. Itoh, Limitations of liver biopsy and non-invasive diagnostic tests for the diagnosis of nonalcoholic fatty liver disease/nonalcoholic steatohepatitis. World journal of gastroenterology: WJG 20, 475 (2014).

12. M. Sheta, E. A. Taha, Y. Lu, T. Eguchi, Extracellular vesicles: new classification and tumor immunosuppression. Biology 12, 110 (2023).

13. G. Van Niel, G. d’Angelo, G. Raposo, Shedding light on the cell biology of extracellular vesicles. Nature reviews Molecular cell biology 19, 213–228 (2018).

14. I. Salido-Guadarrama, S. Romero-Cordoba, O. Peralta-Zaragoza, A. Hidalgo-Miranda, M. Rodriguez-Dorantes, MicroRNAs transported by exosomes in body fluids as mediators of intercellular communication in cancer. OncoTargets and therapy, 1327–1338 (2014).

15. S. Lee, A. M. Gonzalez-Suarez, X. Huang, O. Calvo-Lozano, S. Suvakov, L. M. Lechuga, V. D. Garovic, G. Stybayeva, A. Revzin, Using electrochemical immunoassay in a novel microtiter plate to detect surface markers of preeclampsia on urinary extracellular vesicles. ACS sensors 8, 207–217 (2022).

16. M. C. Ciferri, R. Quarto, R. Tasso, Extracellular vesicles as biomarkers and therapeutic tools: from pre-clinical to clinical applications. Biology 10, 359 (2021).

17. R. Kalluri, V. S. LeBleu, The biology, function, and biomedical applications of exosomes. science 367, eaau6977 (2020).

18. J. Matsuzaki, T. Ochiya, Circulating microRNAs and extracellular vesicles as potential cancer biomarkers: a systematic review. International journal of clinical oncology 22, 413–420 (2017).

19. M. Kornek, M. Lynch, S. H. Mehta, M. Lai, M. Exley, N. H. Afdhal, D. Schuppan, Circulating microparticles as disease-specific biomarkers of severity of inflammation in patients with hepatitis C or nonalcoholic steatohepatitis. Gastroenterology 143, 448–458 (2012).

20. D. Povero, H. Yamashita, W. Ren, M. G. Subramanian, R. P. Myers, A. Eguchi, D. A. Simonetto, Z. D. Goodman, S. A. Harrison, A. J. Sanyal, Characterization and proteome of circulating extracellular vesicles as potential biomarkers for NASH. Hepatology communications 4, 1263–1278 (2020).

21. T. S. Sehrawat, J. P. Arab, M. Liu, P. Amrollahi, M. Wan, J. Fan, Y. Nakao, E. Pose, A. Navarro-Corcuera, D. Dasgupta, Circulating extracellular vesicles carrying sphingolipid cargo for the diagnosis and dynamic risk profiling of alcoholic hepatitis. Hepatology 73, 571–585 (2021).

22. J. Zhou, Z. Wu, J. Hu, D. Yang, X. Chen, Q. Wang, J. Liu, M. Dou, W. Peng, Y. Wu, High-throughput single-EV liquid biopsy: Rapid, simultaneous, and multiplexed detection of nucleic acids, proteins, and their combinations. Science advances 6, eabc1204 (2020).

23. J. Zheng, R. Zhou, B. Wang, C. He, S. Bai, H. Yan, J. Yu, H. Li, B. Peng, Z. Gao, Electrochemical detection of extracellular vesicles for early diagnosis: a focus on disease biomarker analysis. Extracellular Vesicles and Circulating Nucleic Acids 5, 165 (2024).

24. H.-K. Woo, V. Sunkara, J. Park, T.-H. Kim, J.-R. Han, C.-J. Kim, H.-I. Choi, Y.-K. Kim, Y.-K. Cho, Exodisc for rapid, size-selective, and efficient isolation and analysis of nanoscale extracellular vesicles from biological samples. ACS nano 11, 1360–1370 (2017).

25. R. Vaidyanathan, M. Naghibosadat, S. Rauf, D. Korbie, L. G. Carrascosa, M. J. Shiddiky, M. Trau, Detecting exosomes specifically: a multiplexed device based on alternating current electrohydrodynamic induced nanoshearing. Analytical chemistry 86, 11125–11132 (2014).

26. S. Khodashenas, S. Khalili, M. Forouzandeh Moghadam, A cell ELISA based method for exosome detection in diagnostic and therapeutic applications. Biotechnology letters 41, 523–531 (2019).

27. K. Iha, N. Tsurusawa, H.-Y. Tsai, M.-W. Lin, H. Sonoda, S. Watabe, T. Yoshimura, E. Ito, Ultrasensitive ELISA detection of proteins in separated lumen and membrane fractions of cancer cell exosomes. Analytical biochemistry 654, 114831 (2022).

28. S. Lee, D. S. Verkhoturov, M. J. Eller, S. V. Verkhoturov, M. A. Shaw, K. Gwon, Y. Kim, F. Lucien, H. Malhi, A. Revzin, Nanoprojectile secondary ion mass spectrometry enables multiplexed analysis of individual hepatic extracellular vesicles. ACS nano 17, 23584–23594 (2023).

29. E. M. Hassan, M. C. DeRosa, Recent advances in cancer early detection and diagnosis: Role of nucleic acid based aptasensors. TrAC Trends in Analytical Chemistry 124, 115806 (2020).

30. H. Im, H. Shao, Y. I. Park, V. M. Peterson, C. M. Castro, R. Weissleder, H. Lee, Label-free detection and molecular profiling of exosomes with a nano-plasmonic sensor. Nature biotechnology 32, 490–495 (2014).

31. L. Zhu, K. Wang, J. Cui, H. Liu, X. Bu, H. Ma, W. Wang, H. Gong, C. Lausted, L. Hood, Label-free quantitative detection of tumor-derived exosomes through surface plasmon resonance imaging. Analytical chemistry 86, 8857–8864 (2014).

32. C. Liu, X. Zeng, Z. An, Y. Yang, M. Eisenbaum, X. Gu, J. M. Jornet, G. K. Dy, M. E. Reid, Q. Gan, Sensitive detection of exosomal proteins via a compact surface plasmon resonance biosensor for cancer diagnosis. ACS sensors 3, 1471–1479 (2018).

33. P. S. Sfragano, S. Pillozzi, G. Condorelli, I. Palchetti, Practical tips and new trends in electrochemical biosensing of cancer-related extracellular vesicles. Analytical and Bioanalytical Chemistry 415, 1087–1106 (2023).

34. J. Park, J. S. Park, C.-H. Huang, A. Jo, K. Cook, R. Wang, H.-Y. Lin, J. Van Deun, H. Li, J. Min, L. Wang, G. Yoon, B. S. Carter, L. Balaj, G.-S. Choi, C. M. Castro, R. Weissleder, H. Lee, An integrated magneto-electrochemical device for the rapid profiling of tumour extracellular vesicles from blood plasma. Nature biomedical engineering 5, 678–689 (2021).

35. D. G. Mathew, P. Beekman, S. G. Lemay, H. Zuilhof, S. Le Gac, W. G. van der Wiel, Electrochemical detection of tumor-derived extracellular vesicles on nanointerdigitated electrodes. Nano letters 20, 820–828 (2019).

36. N. Gurudatt, H. Gwak, K.-A. Hyun, S.-E. Jeong, K. Lee, S. Park, M. J. Chung, S.-E. Kim, J. H. Jo, H.-I. Jung, Electrochemical detection and analysis of tumor-derived extracellular vesicles to evaluate malignancy of pancreatic cystic neoplasm using integrated microfluidic device. Biosensors and Bioelectronics 226, 115124 (2023).

37. N. Venkatesan, L. C. Doskey, H. Malhi, The role of endoplasmic reticulum in lipotoxicity during metabolic dysfunction–associated steatotic liver disease (MASLD) pathogenesis. The American Journal of Pathology 193, 1887–1899 (2023).

38. L. Zhang, Z. Zhang, C. Li, T. Zhu, J. Gao, H. Zhou, Y. Zheng, Q. Chang, M. Wang, J. Wu, S100A11 promotes liver steatosis via FOXO1-mediated autophagy and lipogenesis. Cellular and molecular gastroenterology and hepatology 11, 697–724 (2021).

39. W. Dornas, D. Glaise, A. Bodin, A. Sharanek, A. Burban, D. Le Guillou, S. Robert, S. Dutertre, C. Aninat, A. Corlu, Endotoxin regulates matrix genes increasing reactive oxygen species generation by intercellular communication between palmitate-treated hepatocyte and stellate cell. Journal of cellular physiology 234, 122–133 (2019).

40. I. Fabregat, J. Moreno-Càceres, A. Sánchez, S. Dooley, B. Dewidar, G. Giannelli, P. Ten Dijke, I. L. Consortium, TGF-β signalling and liver disease. The FEBS journal 283, 2219–2232 (2016).

41. I. D. Vachliotis, S. A. Polyzos, The role of tumor necrosis factor-alpha in the pathogenesis and treatment of nonalcoholic fatty liver disease. Current obesity reports 12, 191–206 (2023).

42. J. Li, H. Liu, A. S. Mauer, F. Lucien, A. Raiter, H. Bandla, T. Mounajjed, Z. Yin, K. J. Glaser, M. Yin, Characterization of cellular sources and circulating levels of extracellular vesicles in a dietary murine model of nonalcoholic steatohepatitis. Hepatology communications 3, 1235–1249 (2019).

43. P. Fattahi, J. M. de Hoyos-Vega, J. H. Choi, C. D. Duffy, A. M. Gonzalez-Suarez, Y. Ishida, K. M. Nguyen, K. Gwon, Q. P. Peterson, T. Saito, Guiding Hepatic Differentiation of Pluripotent Stem Cells Using 3D Microfluidic Co-Cultures with Human Hepatocytes. Cells 12, 1982 (2023).

44. F. Ciregia, M. Bugliani, M. Ronci, L. Giusti, C. Boldrini, M. R. Mazzoni, S. Mossuto, F. Grano, M. Cnop, L. Marselli, Palmitate-induced lipotoxicity alters acetylation of multiple proteins in clonal β cells and human pancreatic islets. Scientific reports 7, 13445 (2017).

45. S. Zhao, J. Jiang, Y. Jing, W. Liu, X. Yang, X. Hou, L. Gao, L. Wei, The concentration of tumor necrosis factor-α determines its protective or damaging effect on liver injury by regulating Yap activity. Cell death & disease 11, 70 (2020).

46. A. Dropmann, T. Dediulia, K. Breitkopf-Heinlein, H. Korhonen, M. Janicot, S. N. Weber, M. Thomas, A. Piiper, E. Bertran, I. Fabregat, TGF-β1 and TGF-β2 abundance in liver diseases of mice and men. Oncotarget 7, 19499 (2016).

47. J. A. Welsh, D. C. Goberdhan, L. O’Driscoll, E. I. Buzas, C. Blenkiron, B. Bussolati, H. Cai, D. Di Vizio, T. A. Driedonks, U. Erdbrügger, Minimal information for studies of extracellular vesicles (MISEV2023): From basic to advanced approaches. Journal of extracellular vesicles 13, e12404 (2024).

48. H. Malhi, S. F. Bronk, N. W. Werneburg, G. J. Gores, Free fatty acids induce JNK-dependent hepatocyte lipoapoptosis. Journal of Biological Chemistry 281, 12093–12101 (2006).

49. A. M. Allen, V. H. Shah, T. M. Therneau, S. K. Venkatesh, T. Mounajjed, J. J. Larson, K. C. Mara, P. J. Schulte, T. A. Kellogg, M. L. Kendrick, The role of 3D-MRE in the diagnosis of NASH in obese patients undergoing bariatric surgery. Hepatology (Baltimore, Md.) 71, 510 (2019).

50. A. M. Allen, College of Medicine-Mayo Clinic, (2023).

51. M. Charlton, K. Viker, A. Krishnan, S. Sanderson, B. Veldt, A. Kaalsbeek, M. Kendrick, G. Thompson, F. Que, J. Swain, Differential expression of lumican and fatty acid binding protein-1: new insights into the histologic spectrum of nonalcoholic fatty liver disease. Hepatology 49, 1375–1384 (2009).

52. A. H. Ali, B. D. Juran, E. M. Schlicht, J. K. Bianchi, B. M. McCauley, E. J. Atkinson, K. N. Lazaridis, The PSC scientific community resource: an asset for multi-omics interrogation of primary sclerosing cholangitis. BMC gastroenterology 21, 353 (2021).

53. J. Ludwig, E. Dickson, G. S. A. McDonald, Staging of chronic nonsuppurative destructive cholangitis (syndrome of primary biliary cirrhosis). Virchows Archiv A 379, 103–112 (1978).

54. S. Lee, B. P. Crulhas, S. Suvakov, S. V. Verkhoturov, D. S. Verkhoturov, M. J. Eller, H. Malhi, V. D. Garovic, E. A. Schweikert, G. Stybayeva, Nanoparticle-enabled multiplexed electrochemical immunoassay for detection of surface proteins on extracellular vesicles. ACS applied materials & interfaces 13, 52321–52332 (2021).

