## Supplemental information for "Electrochemical immunosensor-enabled liquid biopsy of extracellular vesicles for diagnosis of metabolic dysfunction-associated steatohepatitis"

### MATERIALS AND METHODS

#### Materials

4-morpholineethanesulfonic acid (MES), 11-mercaptoundecanoic acid (MUA), 1-ethyl-3-(3-dimethylaminopropyl carbodiimide) (EDC), N-hydroxy-succinimide (NHS), Casein, Bovine serum Albumin (BSA), Pluronic F-127, Palmitate (PA), and $Pb(NO_3)_2$ were purchased from Sigma-Aldrich (St. Louis, MO, USA). Blocker$^{TM}$ Casein in PBS and SuperBlock$^{TM}$ Blocking buffer were purchased from Thermo Fisher Scientific (Waltham, MA, USA). Polydimethylsiloxane (PDMS, Sylgard 184 Silicone Elastomer Kit) and SU-8 2100 were purchased from Ellsworth (Minneapolis, MN, USA) and Kayaku Advanced Materials (Westborough, MA, USA), respectively. Dulbecco's modified Eagle's medium (DMEM) and Dulbecco's Phosphate-buffered Saline (DPBS) was purchased from Corning (Corning, NY, USA). Fetal bovine serum (FBS) and penicillin/streptomycin (P/S) were purchased from Gibco (Grand Island, NY, USA). Exosome-depleted FBS was from Captivate Bio (Watertown, MA, USA). Primers for RT-PCR were from IDT (Coralville, IA, USA) and Sigma-Aldrich. Paraformaldehyde (PFA) was purchased from Election Microscopy Sciences (Hatfield, PA, USA).

Antibodies for rabbit anti-human ASGR2, chicken anti-human TGF-β1, goat anti-human TNF-α and goat anti-human S100A11 were purchased from R&D systems (Minneapolis, MN, USA). Mouse anti-human CD63 and mouse IgG isotype control were purchased from BD Biosciences (San Jose, CA, USA). Rabbit anti-human CYP2E1 antibody was purchased from CYP450-GP (Vista, CA, USA). Rabbit IgG isotype control was purchased from Thermo Fisher Scientific (Waltham, MA, USA). Lipid droplet stain (Lipi-Red) was purchased from Dojindo (Mashiki, Tabaru, Japan). Rabbit anti-human S100A11 polyclonal antibody was purchased from Proteintech (Rosemont, IL, USA). Goat anti-human albumin antibody was purchased from Bethyl Laboratories (Montgomery, TX, USA). Alexa Fluor 546 (donkey anti-chicken IgY and donkey anti-rabbit IgG), Donkey anti-goat IgG Alexa Fluor 488 and Donkey anti-goat IgG Alexa Fluor 647 were purchased from Life Technologies (Carlsbad, CA, USA). DAPI was purchased from BD Bioscience (San Jose, CA, USA).

NHS-activated AuNPs (20 nm) were purchased from CytoDiagnostics (Tulsa, Oklahoma, USA). Glass slides sputtering of 10nm Cr and 100nm Au were fabricated from LGA Thin Films (Lance Goddard Associates, Santa Clara, CA). Ag/AgCl ink was purchased from CH Instruments (Bee Cave, TX, USA). Poly(dimethylsiloxane) (PDMS) base and curing agent kit (Sylgard184) were purchased from Ellsworth Adhesives (Minneapolis, MN, USA).

#### Cell culture

Human hepatocytes were a kind gift from Prof. Takeshi Saito at the University of South California and Phoenix Bio Ltd. These cells were isolated from chimeric mice with humanized livers (cDNA- uPA+/−/SCID (uPA+/wt: B6;129SvEv- Plau, SCID: C.B- 17/Icr- scid/scid Jcl)) using standard cannulation and collagenase perfusion protocol (*1*). Human hepatocytes were cultured in DMEM supplemented with 20 mM HEPES, 10% FBS, 100 µg/mL streptomycin, 100 U/mL penicillin, 15 µg/mL L-Proline, 50 nM Dexamethasone, 0.25 µg/mL recombinant human insulin 5 µg/mL EGF, 0.1 mM L-ascorbic acid 2-phosphate and 2% DMSO at 37°C in a humidified 5% $CO_2$ atmosphere. All the cell lines were confirmed to be free of mycoplasma contamination prior to use.

**Cell characterization**

For immunofluorescence staining, cells were fixed with 4% PFA overnight at 4ºC. Then, cells were immersed in 0.1% Triton X-100 for permeabilization for 20 min, rinsed thoroughly and then incubated with primary antibodies (albumin, TGF-β1, TNF-α, and S100A11) in 1% BSA-PBS for 1.5 h. Afterwards, cells were incubated with either fluorescently-labeled secondary antibodies (2 µg/mL), DAPI, phalloidin or lipid droplet staining solution (Lipi-Red) in 1% BSA-PBS solution for another 1 h in dark. The imaging was performed using inverted fluorescence microscope (Olympus IX83).

For RT-PCR, cells were lysed using cell lysis buffer and dissolved completely using vortex for 10 seconds. Then, total RNA was extracted using an RNA lysis kit (Qiagen; Valencia, CA, USA) according to the manufacturer's protocol. Approximately 10-50 ng/µL was used for synthesis of cDNA using the reverse transcription kit (Roche). Gene expression was performed using a QuantStudioTM 5 System (Thermo Fisher Scientific) with SYBR Green and was normalized to glyceraldehyde 3-phosphate dehydrogenase (GAPDH). The amplification procedure for RT-PCR consists of 40 cycles of denaturation at 95°C for 5 s, annealing at 55°C for 15 s, and extension at 69°C for 20 s. The final analysis was operated based on the threshold cycles using the ΔΔCT method. The primers for PCR are listed in Table S1.

Table S1. List on sequences of primers for RT-PCR analysis

| Gene | Forward | Reverse |
|---|---|---|
| **GAPDH** | 5’-AGACAGCCGCATCCTCTTGT-3’ | 5’-CTTGCCGTGGGTAGAGTCAT-3’ |
| **TGF-β1** | 5’-CCTGGAAAGGGCTCAACAC-3’ | 5’-CGATTCTTCTCTGTGGAGCTG-3’ |
| **TNF-α** | 5’-AGGCAGTCAGATCATCTTC-3’ | 5’-TTATCTCTCAGCTCCACG-3’ |
| **S100A11** | 5’-CCAGAAGTATGCTGGAAAGGATG-3’ | 5’-CATCATGCGGTCAAGGACACCA-3’ |
| **CYP2E1** | 5’-GACACCATTTTCAGAGGATAC-3’ | 5’-TTCATTCAGGAAGTGTTCTG-3’ |
| **Albumin** | 5’-AGCCTACCATGAGAATAATAG-3’ | 5’-TTGAAGCACAGAGAAAAGAG-3’ |

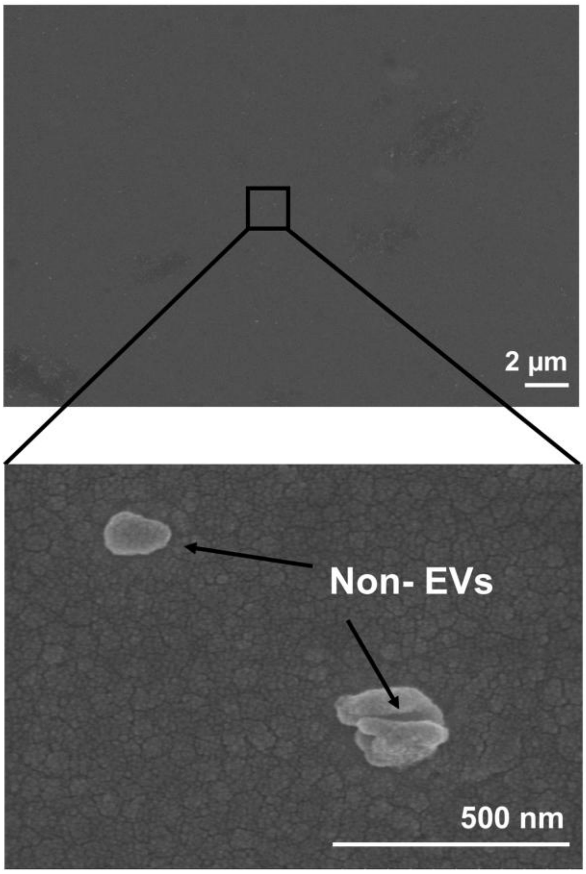


Figure S1. SEM image of Au substrate that was modified with IgG isotype control Abs and incubated with $5{\times}10^9$ EVs/mL for 1 h.

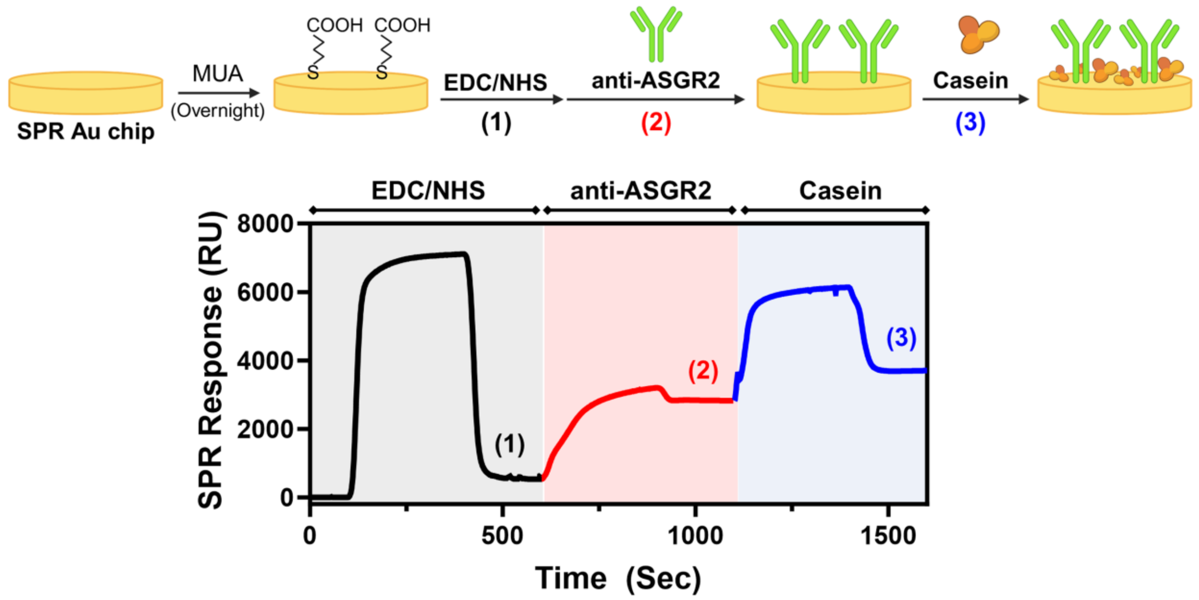


Figure S2. Cartoon illustrating steps in the assembly of the biorecognition layer on the Au substrate of SPR. SPR sensogram shows changes in signal due to functionalization steps outlined in the cartoon.

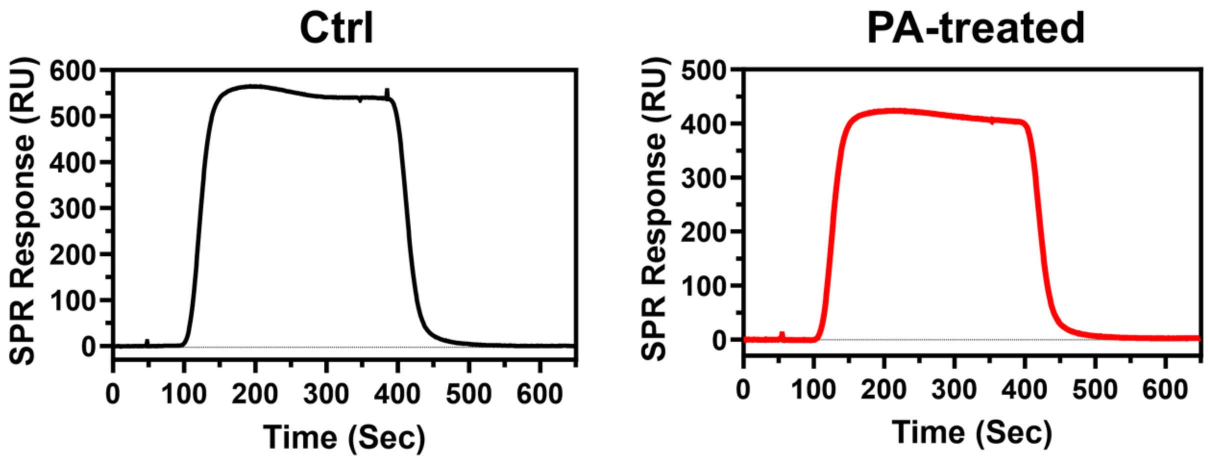


Figure S3. SPR curves on adsorption IgG isotype control on human hepatocyte control and 0.4 mM PA exposed EVs.

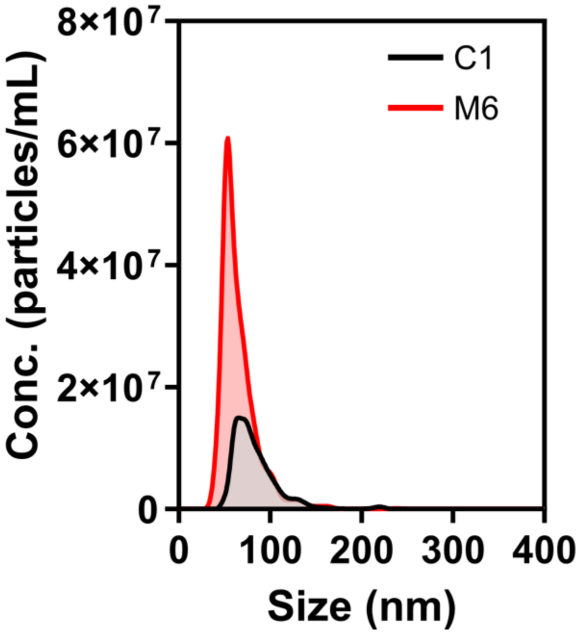


Figure S4. Size distribution of EVs from selected healthy control (Ctrl 1) and MASH (MASH 6) samples.

Table S2. Summary of patient-derived plasma EV concentration and size for each cohort.

| | **Ctrl (n=8)** | **MASH (n=10)** |
|---|---|---|
| **Concentration ($10^{10}$ particles/mL)** | 3.77 ± 1.67 | 8.41 ± 2.04 |
| **Size (nm)** | 73.15 ± 4.86 | 66.82 ± 9.15 |

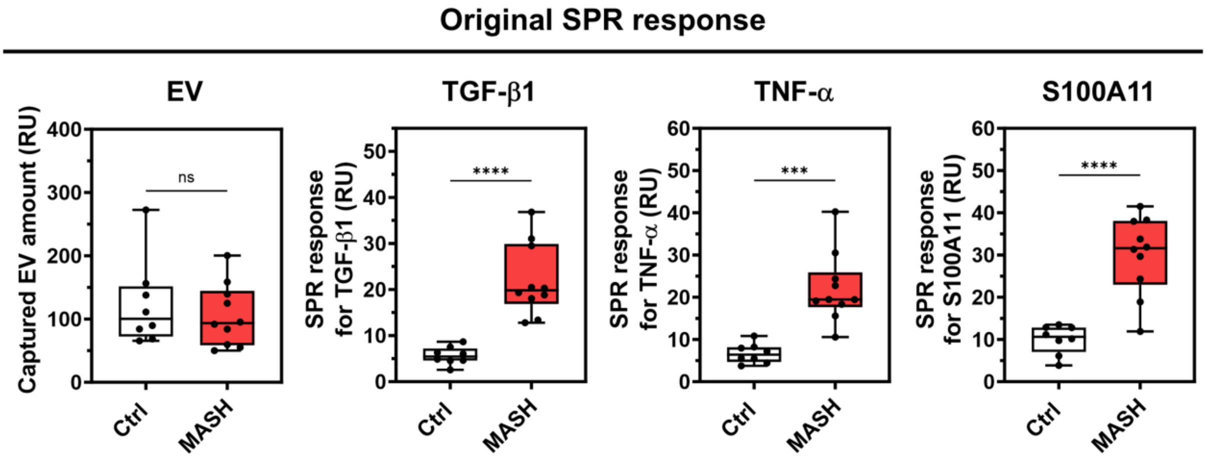


Figure S5. Original SPR response (RU) showing the captured amount of plasma EV and the detection level of TGF-β1, TNF-α and S100A11 on EV.

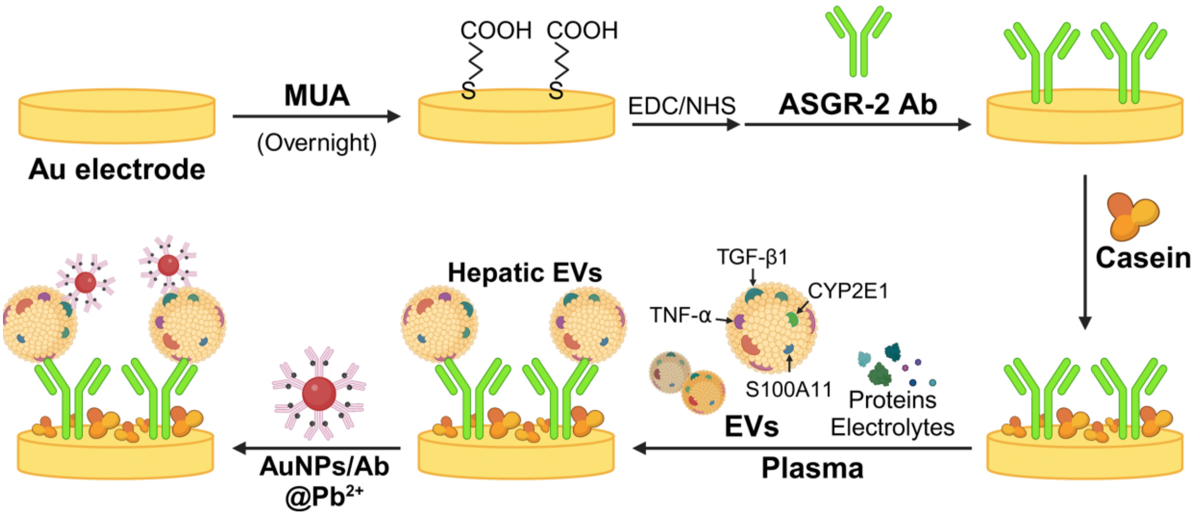


Figure S6. Sequential steps for electrode functionalization with capture antibodies, EV capture, and labeling with immunoprobes.

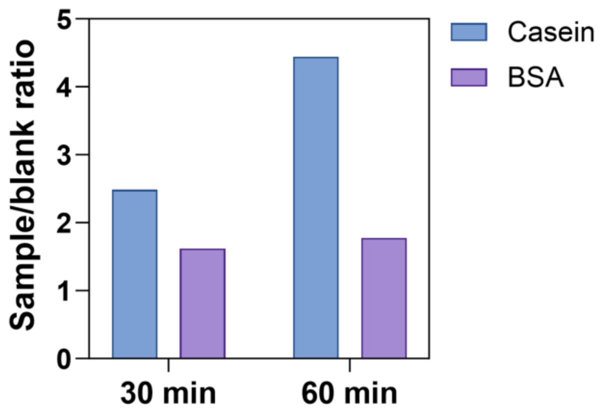


Figure S7. Optimization for blocking reagents including BSA and Casein. The sample/blank ratio is calculated by dividing electrochemical response of MASH to Control.

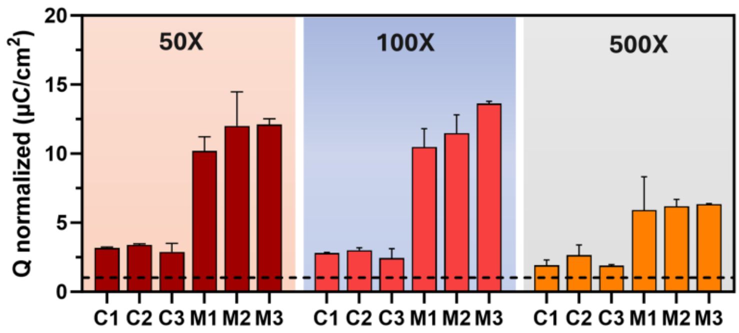


Figure S8. Optimization for plasma sample dilution ratio. The plasma is diluted with Superblock and conducted under same experimental conditions (Healthy control n = 3, MASH n = 3).

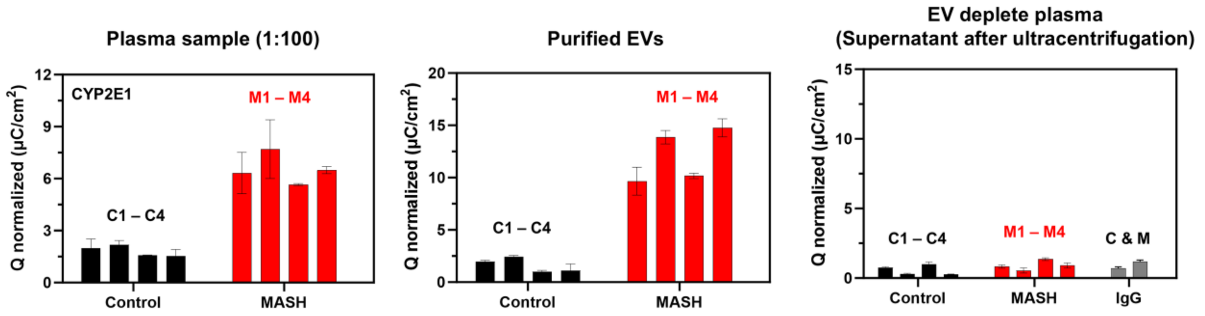


Figure S9. Performance of the electrochemical microtiter plate with different types of samples including purified EVs, EV-depleted plasma and diluted plasma. Anti-ASGR-2 is used as the capture antibodies and AuNPs/CYP2E1@$Pb^{2+}$ is used as the target immunoprobe.

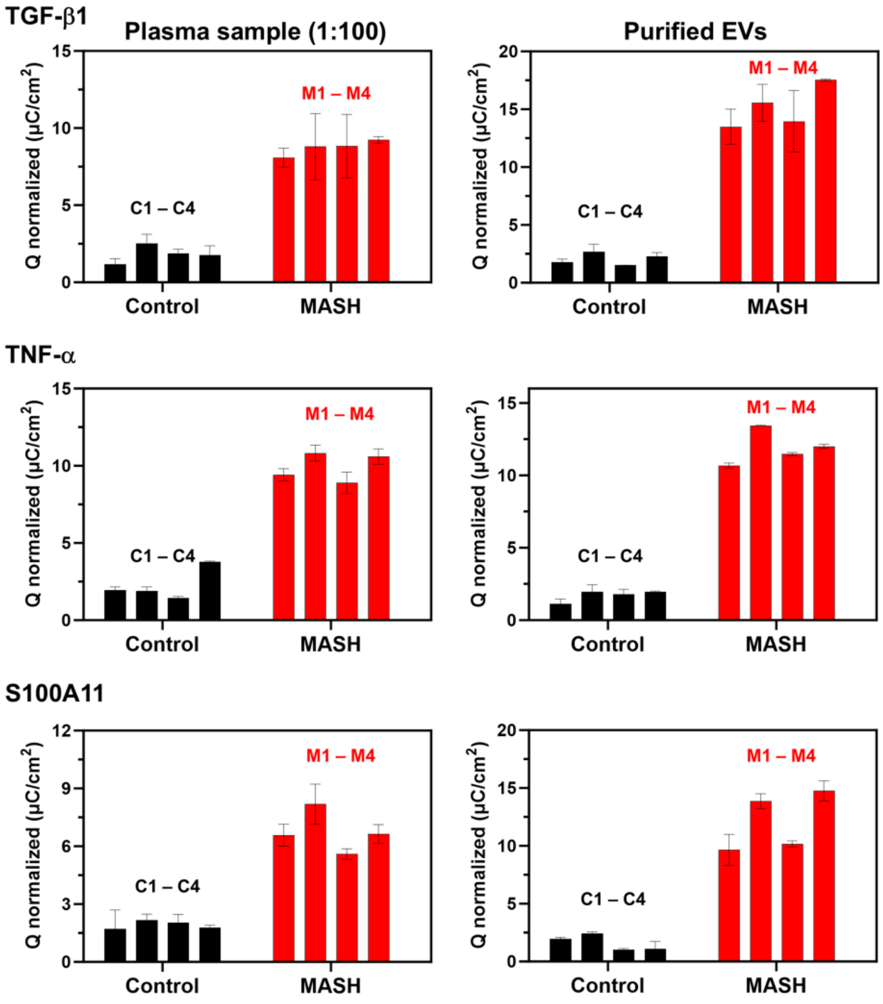

Figure S10. Comparison of electrochemical signals between purified EVs and original plasma samples for the detection of TGF-β1, TNF-α, and S100A11 in MASH and healthy control cohorts. Plasma samples were diluted (1:100) before measurement. Electrochemical signals were normalized and presented as mean ± SD. C1–C4 represent individual control samples, while M1–M4 represent MASH patient samples.

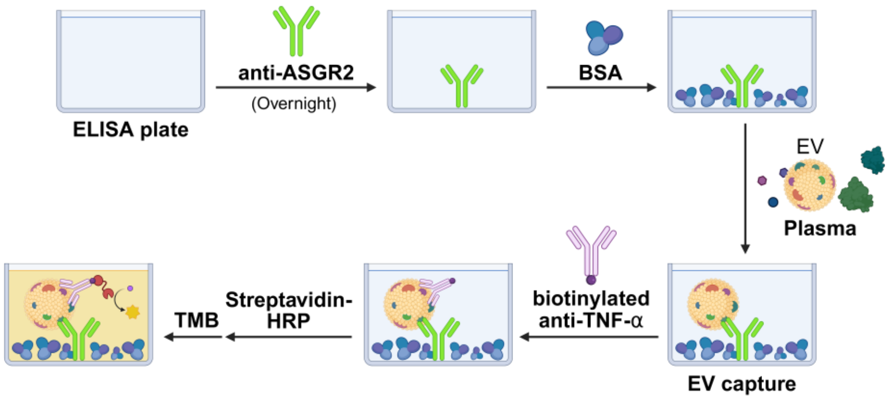


Figure S11. Illustration of workflow on customized colorimetric ELISA for determining TNF-α expression on hepatic EVs in plasma.

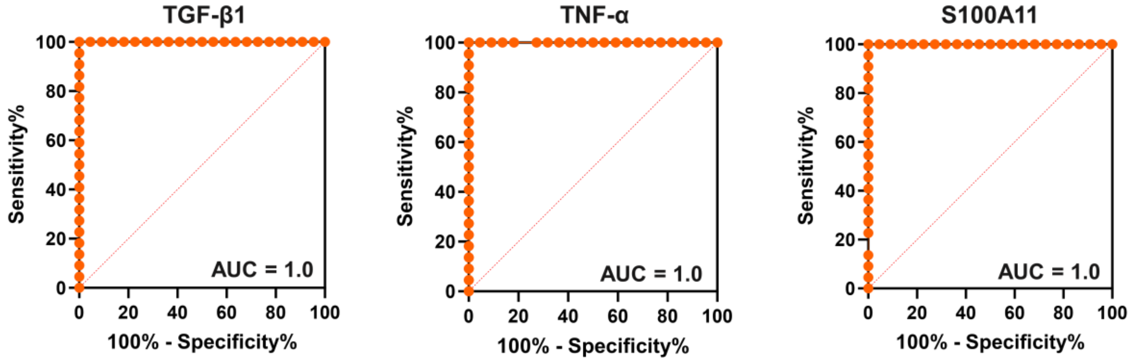


Figure S12. ROC analysis of individual MASH-associated EV biomarkers. ROC curves for TGF-β1, TNF-α, and S100A11 measured by the electrochemical immunoassay in the training cohort. All three biomarkers achieved an AUC of 1.0, indicating complete separation between MASH and healthy control samples within this cohort.

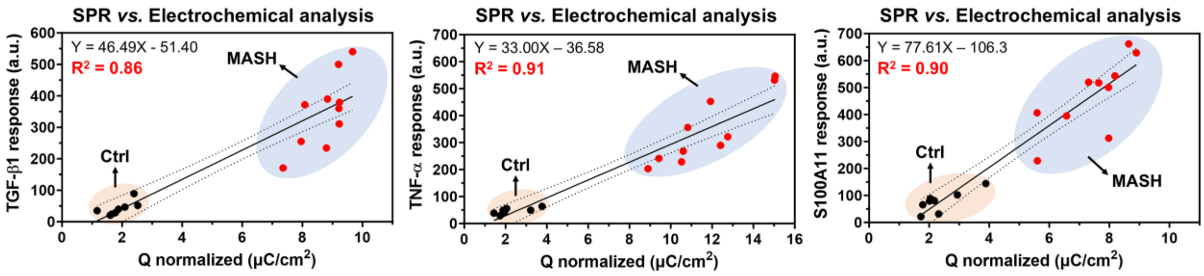


Figure S13. Correlation between electrochemical measurements and SPR analysis. Linear correlation analysis comparing electrochemical signal intensities obtained from plasma samples with SPR responses measured from corresponding purified EV samples. Strong correlations were observed for TGF-β1 ($R^2$ = 0.86), TNF-α ($R^2$ = 0.91), and S100A11 ($R^2$ = 0.90), supporting the quantitative reliability of the electrochemical platform.

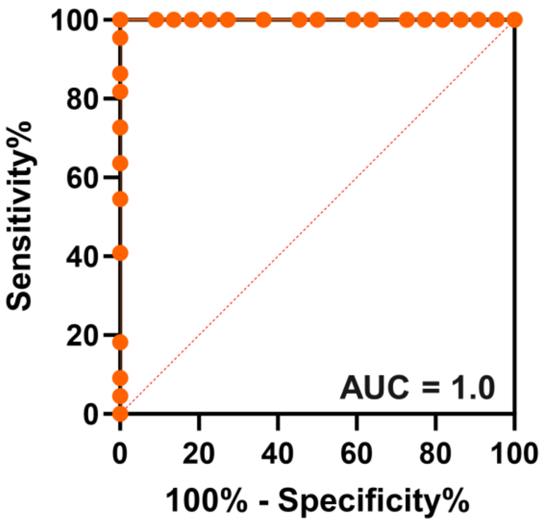


Figure S14. ROC curve for the multi-marker composite score ($EV_{MASH}$) derived from combined TGF-β1, TNF-α, and S100A11 signals in the training cohort. The composite score achieved an AUC of 1.0, comparable to individual biomarkers.

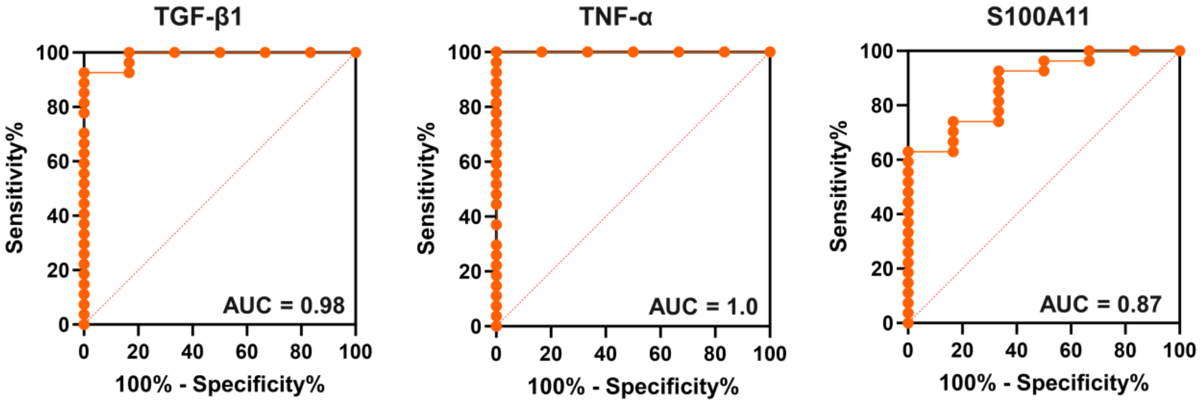


Figure S15. ROC analysis of individual MASH-associated EV biomarkers in validation cohort. ROC curves for TGF-β1, TNF-α, and S100A11 measured by the electrochemical immunoassay between MASH and non-MASH patient groups.

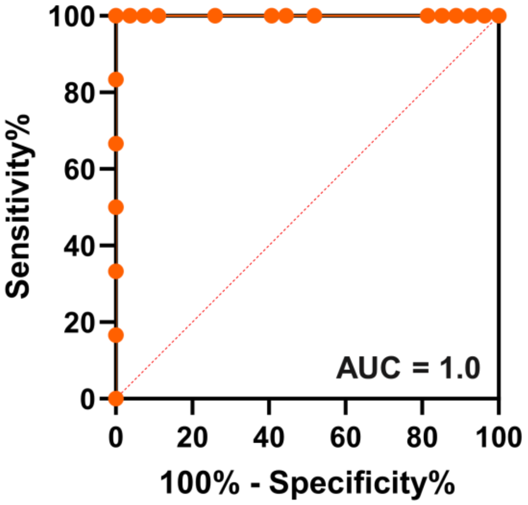


Figure S16. ROC curve for the multi-marker composite score ($EV_{MASH}$) derived from combined TGF-β1, TNF-α, and S100A11 signals in the training cohort. The composite score achieved an AUC of 1.0 outperforming individual biomarkers.

Table S3. Summary of MASH diagnostic statistics for EV markers.

| | Training cohort (n = 44) | | | |
|---|---|---|---|---|
| **Cut-off** | **Sensitivity (%)** | **Specificity (%)** | **Accuracy (%)** | **Youden index** |
| > 7.550 | 100 | 50 | 75.00 | 0.5 |
| > 7.850 | 100 | 54.55 | 77.27 | 0.5455 |
| > 8.150 | 100 | 63.64 | 81.82 | 0.6364 |
| > 8.500 | 100 | 72.73 | 86.36 | 0.7273 |
| > 8.650 | 100 | 77.27 | 88.64 | 0.7727 |
| > 8.900 | 100 | 81.82 | 90.91 | 0.8182 |
| > 9.150 | 100 | 86.36 | 93.18 | 0.8636 |
| > 9.600 | 100 | 90.91 | 95.45 | 0.9091 |
| **> 15.00** | **100** | **100** | **100.00** | **1** |
| > 20.50 | 95.45 | 100 | 97.73 | 0.9545 |
| > 21.50 | 86.36 | 100 | 93.18 | 0.8636 |
| > 22.50 | 81.82 | 100 | 90.91 | 0.8182 |
| > 23.50 | 72.73 | 100 | 86.36 | 0.7273 |
| > 24.50 | 63.64 | 100 | 81.82 | 0.6364 |
| > 25.50 | 54.55 | 100 | 77.27 | 0.5455 |

Table S4. Histological grading of MASH patients based on liver biopsy evaluation.

| MASH | | | | |
|---|---|---|---|---|
| No. | Stage of fibrosis | Steatosis grade | Lobular inflammation grade | Group |
| 1 | 1 | 2 | 2 | MASH + F 0-1 |
| 2 | 1 | 2 | 1 | MASH + F 0-1 |
| 3 | 1 | 2 | 1 | MASH + F 0-1 |
| 4 | 2 | 2 | 1 | MASH + F 2-4 |
| 5 | 3 | 3 | 1 | MASH + F 2-4 |
| 6 | 3 | 1 | 2 | MASH + F 2-4 |
| 7 | 2 | 2 | 1 | MASH + F 2-4 |
| 8 | 2 | 2 | 1 | MASH + F 2-4 |
| 9 | 3 | 1 | 2 | MASH + F 2-4 |
| 10 | 2 | 1 | 1 | MASH + F 2-4 |
| 11 | 1 | 1 | 2 | MASH + F 0-1 |
| 12 | 1 | 1 | 2 | MASH + F 0-1 |
| 13 | 1 | 1 | 2 | MASH + F 0-1 |
| 14 | 1 | 1 | 2 | MASH + F 0-1 |
| 15 | 1 | 1 | 2 | MASH + F 0-1 |
| 16 | 1 | 2 | 2 | MASH + F 0-1 |
| 17 | 1 | 1 | 2 | MASH + F 0-1 |
| 18 | 1 | 1 | 2 | MASH + F 0-1 |
| 19 | 2 | 1 | 1 | MASH + F 2-4 |
| 20 | 2 | 2 | 1 | MASH + F 2-4 |
| 21 | 2 | 2 | 2 | MASH + F 2-4 |
| 22 | 2 | 1 | 1 | MASH + F 2-4 |
| 23 | 3 | 1 | 1 | MASH + F 2-4 |
| 24 | 2 | 1 | 2 | MASH + F 2-4 |
| 25 | 2 | 0 | 1 | MASH + F 2-4 |
| 26 | 2 | 0 | 1 | MASH + F 2-4 |
| 27 | 3 | 2 | 1 | MASH + F 2-4 |
| 28 | 3 | 2 | 1 | MASH + F 2-4 |

Table S5. Histological grading of Simple Steatosis patients based on liver biopsy evaluation.

| | Simple Steatosis | | | |
|---|---|---|---|---|
| **No.** | **Stage of fibrosis** | **Steatosis grade** | **Lobular inflammation grade** | **Group** |
| 1 | 0 | 2 | 0 | Simple steatosis |
| 2 | 0 | 2 | 0 | Simple steatosis |
| 3 | 0 | 1 | 0 | Simple steatosis |
| 4 | 0 | 1 | 0 | Simple steatosis |
| 5 | 0 | 1 | 0 | Simple steatosis |
| 6 | 0 | 1 | 0 | Simple steatosis |

Table S6. Histological grading of Obese normal patients based on liver biopsy evaluation.

| | Obese Normal | | | |
|---|---|---|---|---|
| **No.** | **Stage of fibrosis** | **Steatosis grade** | **Lobular inflammation grade** | **Group** |
| 1 | 0 | 0 | 0 | Obese normal |
| 2 | 0 | 0 | 0 | Obese normal |
| 3 | 0 | 0 | 0 | Obese normal |
| 4 | 0 | 0 | 0 | Obese normal |

Table S7. Histological grading of Cholestatic liver disease patients based on liver biopsy evaluation.

Table S7. Histological grading of Cholestatic liver disease patients based on liver biopsy evaluation.

| | Cholestatic Liver Diseases | | |
|---|---|---|---|
| **No.** | **PSC stage** | **PBC stage** | **Group** |
| 1 | | 1 | PBC |
| 2 | | 2 | PBC |
| 3 | | 1 | PBC |
| 4 | | 0 | PBC |
| 5 | | 1 | PBC |
| 6 | | 4 | PBC |
| 7 | | 4 | PBC |
| 8 | | 4 | PBC |
| 9 | 1 | | PSC |
| 10 | 2 | | PSC |
| 11 | 1.5 | | PSC |
| 12 | 1 | | PSC |
| 13 | 1 | | PSC |
| 14 | 0.5 | | PSC |
| 15 | 3 | | PSC |
| 16 | 3 | | PSC |

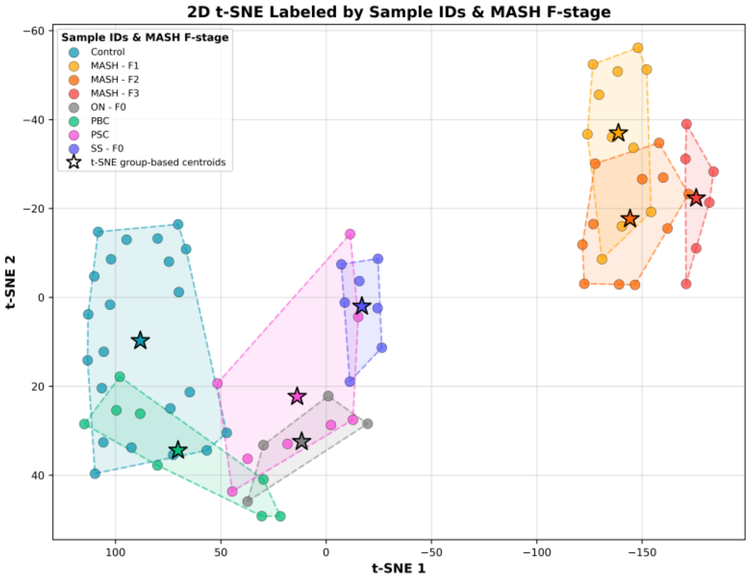


Figure S17. t-distributed stochastic neighbor embedding (t-SNE) analysis of electrochemical signals demonstrating clear clustering trends distinguishing fibrosis stage 3 MASH samples fibrosis stage 1 and 2 MASH samples. (n = 77, features = 3, perplexity = 10, learning rate = 200). The stars denote the centroids of each disease group and MASH fibrosis stage.

Table S8. Evaluation of t-SNE clustering with a silhouette score for each clinical group.

| | **Silhouette Scores** | | | | |
|---|---|---|---|---|---|
| **Sample ID (n)** | **Mean** | **Median** | **SD** | **Min** | **Max** |
| Control (22) | 0.205 | 0.318 | 0.23 | -0.271 | 0.439 |
| MASH (28) | 0.757 | 0.766 | 0.05 | 0.618 | 0.825 |
| ON (4) | -0.139 | -0.093 | 0.16 | -0.354 | -0.016 |
| PBC (8) | -0.300 | -0.293 | 0.18 | -0.506 | 0.010 |
| PSC (8) | -0.367 | -0.295 | 0.21 | -0.736 | -0.133 |
| SS (7) | 0.581 | 0.613 | 0.15 | 0.257 | 0.695 |
| **Global Silhouette Score (Mean)** | | | 0.31 | | |

The table details the mean, median, standard deviation, and range of Silhouette coefficients calculated directly from the 2D t-SNE embedding. Scores range from -1 to +1, where positive values denote distinct visual cluster separation and negative values indicate spatial overlap within the projection.

Table S9. Evaluation of t-SNE clustering with a silhouette score for each clinical group with the MASH group separated into three separate groups based on fibrosis stage.

| | Silhouette Scores | | | | |
|---|---|---|---|---|---|
| **Sample ID (n)** | **Mean** | **Median** | **SD** | **Min** | **Max** |
| Control (22) | 0.20 | 0.32 | 0.23 | -0.271 | 0.439 |
| MASH F1 (11) | 0.16 | 0.32 | 0.31 | -0.340 | 0.439 |
| MASH F2 (11) | -0.04 | -0.01 | 0.28 | -0.601 | 0.309 |
| MASH F3 (6) | 0.47 | 0.47 | 0.12 | 0.284 | 0.612 |
| ON (4) | -0.14 | -0.09 | 0.16 | -0.354 | -0.016 |
| PBC (8) | -0.30 | -0.29 | 0.18 | -0.506 | 0.010 |
| PSC (8) | -0.37 | -0.30 | 0.21 | -0.736 | -0.133 |
| SS (7) | 0.58 | 0.61 | 0.15 | 0.257 | 0.695 |
| **Global Silhouette Score (Mean)** | | | 0.09 | | |

**Reference**


1. S. Lee, D. S. Verkhoturov, M. J. Eller, S. V. Verkhoturov, M. A. Shaw, K. Gwon, Y. Kim, F. Lucien, H. Malhi, A. Revzin, Nanoprojectile secondary ion mass spectrometry enables multiplexed analysis of individual hepatic extracellular vesicles. *ACS nano* **17**, 23584–23594 (2023).
